# A genome-wide association study in Peruvians suggests new risk loci for Alzheimer Disease

**DOI:** 10.1101/2023.11.29.23299201

**Authors:** Bilcag Akgun, Mario Cornejo-Olivas, Nilton Custodio, Pedro R Mena, Marcio Soto-Añari, Rosa Montesinos, Larry D Adams, Katrina Celis, Zikun Yang, Basilio C Huaman, Dolly Reyes-Dumeyer, Joe Rivero, Patrice G Whitehead, Kara L Hamilton-Nelson, Liyong Wang, Derek M Dykxhoorn, Julia Rios-Pinto, Angel Medina-Colque, Clifton L Dalgard, Eden R Martin, Ivan Cornejo-Herrera, Maryenela Illanes-Manrique, Edward Ochoa-Valle, Elison Sarapura-Castro, Aura M Ramirez, Sofia Moura, Wanying Xu, Koni K Mejía, Karina Milla-Neyra, Sheila Castro-Suarez, Alzheimer’s Disease Sequencing Project (ADSP), Fulai Jin, Anthony J Griswold, Gary W Beecham, Rosario Isasi, Katalina F McInerney, Michael L Cuccaro, Jeffery M Vance, Farid Rajabli, Margaret A Pericak-Vance, Giuseppe Tosto

## Abstract

Increasing ethnic/ancestral diversity in genetic studies is critical for defining the genetic architecture of Alzheimer disease (AD). Amerindian (AI) populations are substantially underrepresented in AD genetic studies. The Peruvian population, with up to ∼80% of AI ancestry, provides a unique opportunity to assess the role of AI ancestry in AD. We aimed to conduct a genome-wide association study and comprehensive analyzes to identify novel ancestry-specific AD susceptibility loci and characterize the known AD genetic risk loci in the Peruvian population. 528 individuals were included in these analyses (175 AD, 353 cognitively unimpaired). Array genotype was imputed to the NHLBI TOPMedv5 haplotype reference panel. We used a generalized linear mixed model adjusting for sex, age, and population substructure (model-1), and adjusting for *APOE-ε4* allele dosage (model-2). Both models included a genetic relationship matrix as a random effect to account for cryptic relatedness. To determine if the associations were ancestry-specific, we further explored interactions between top genetic variants and local ancestry. Single nuclei ATACseq and RNAseq, and Hi-C analysis was performed to identify potential interactions with the associated locus. We identified a novel genome-wide significant signal (rs2625222; *P*=3.5×10^-8^) within the Neurofascin gene (*NFASC)* on chromosome 1. Local ancestry approach estimated both European and African ancestral backgrounds at the *NFASC* locus. Functional studies found that rs2625222 lies within an open ATAC peak and is expressed in oligodendrocytes and neurons. Hi-C studies revealed strong loops between rs2625222 region and the promoter of *NFASC* in neurons and oligodendrocytes. We also replicated three known AD loci: *APOE*, *TREML2*, and *CLU*.

This study identified a novel locus associated with risk for AD in the Peruvian population. Functional studies strongly support that *NFASC* is the targeted gene of this risk association. These findings emphasize the importance of including diverse populations in genetic studies of AD.

**Author summary:** Alzheimer Disease (AD), the most common type of dementia in older adults, has a complex etiology with a strong genetic predisposition. Despite Genome-Wide Association Studies (GWAS) identifying over 75 loci associated with AD, these have predominantly focused on the non-Hispanic White population. Genetic studies of diverse populations, such as the Peruvian population with its significant Amerindian ancestral background, are crucial for identifying ancestry-specific genetic risk and protective loci associated with AD. Our objective in this study was to conduct a GWAS and comprehensive analyses of 528 Peruvian individuals to identify new ancestry-specific AD risk loci. In this study, we identified a genome-wide significant novel signal within the *NFASC* gene, which has a crucial role in central and peripheral nervous systems, on chromosome 1. This locus showed both European and African ancestral backgrounds. Our functional analyses strongly support that the *NFASC* gene itself is the primary gene tagged by the risk-associated signal. These findings emphasize the inclusion of admixed populations in genetic studies provides an important opportunity to assess the role of different ancestries in AD.

## Introduction

Alzheimer Disease (AD), the most common type of dementia in the older adults worldwide, accounts for an estimated more than 60% of all dementia cases [1]. The prevalence of AD increases with age, affecting more than a third of individuals above age 85 [2]. The etiology of AD is complex with a strong genetic predisposition [3, 4]. Genome-wide association studies (GWAS) have identified more than 75 loci associated with AD to date[5]. However, these studies have primarily focused on non-Hispanic White (NHW) populations [6, 7]. Research into AD genetics across diverse populations reveals a partial overlap of genetic risk and protective loci among different ancestral groups, while also showing differences in effect sizes and specific genetic variants associated with AD [8–11]. Including diverse populations in AD genetic studies emphasizes the importance of identifying ancestry-specific loci, the importance of local ancestry, as well as generalizing risk and protective loci across ancestral populations [12]. Notably, Amerindian (AI) ancestry populations are among the least represented in AD genetic studies [7], highlighting the necessity of extending AD genetic studies to populations with a high proportion of AI ancestral background. This is essential for ensuring a more comprehensive understanding of AD genetic architecture and fostering the development of precision medicine.

The diverse and multicultural Peruvian population, characterized by its substantial AI ancestral background offers a unique opportunity to identify ancestry-specific genetic risk and protective loci associated with AD and generalize known AD loci to the Peruvian population. The estimated dementia prevalence among Peruvians with age over 65 is 7% [13]. The Peruvian population is an admixed population due to migration events during and after the Inca Empire and Spanish colonization [14]. Peruvians have on average 80% of AI ancestral background, followed by European (EU), East Asian (EA) and African (AF) [14]. The relationship between AD risk loci other than *APOE* and the risk of AD in populations of AI descent is not clear.

In this study, we aimed to conduct a genome-wide association study (GWAS) and comprehensive analyzes to identify novel ancestry-specific AD susceptibility loci and characterize the known AD genetic risk loci in the Peruvian population using functional approaches.

## Results

### Association analysis

A total of 175 individuals (122 female [69.7%]) with AD and 353 cognitively unimpaired (222 female [62.9%]) were analyzed (Table 1). There was no evidence for genomic inflation (model 1: λ = 1.055; model 2: λ = 1.043) (S1 Fig.). A novel locus, *NFASC* on chromosome 1, reached genome-wide significance (P ≤ 5×10^-8^) in both models (Table 2, Fig. 1). Two novel loci reached suggestive significance at P ≤5×10^−7^: *STK32A* on chromosome 5, and *RP11-663N22.1* on chromosome 17 (Table 2, Fig. 1).

**Figure 1.**
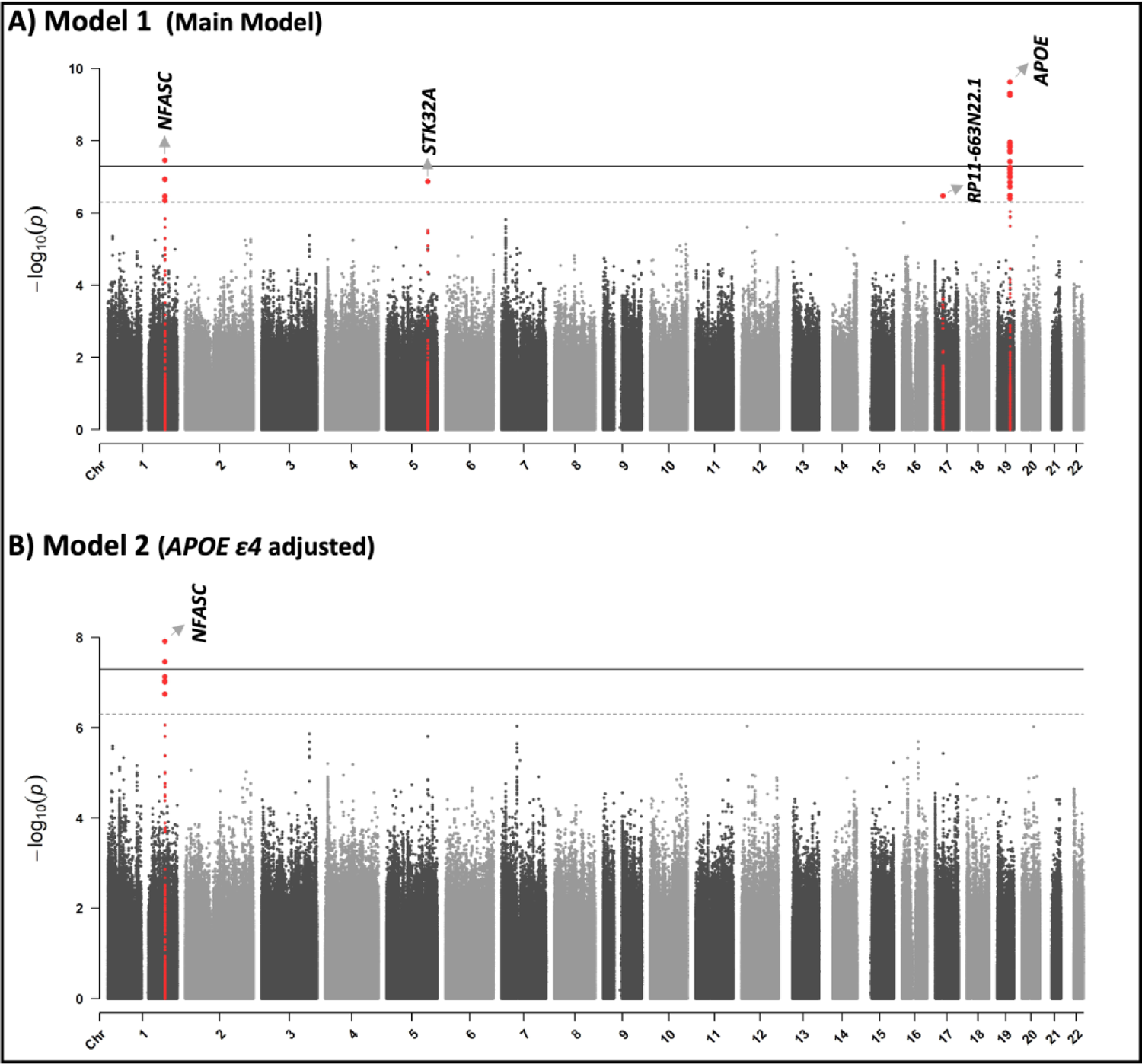
Manhattan plots of Single Variant Analysis.

**Table 1.**
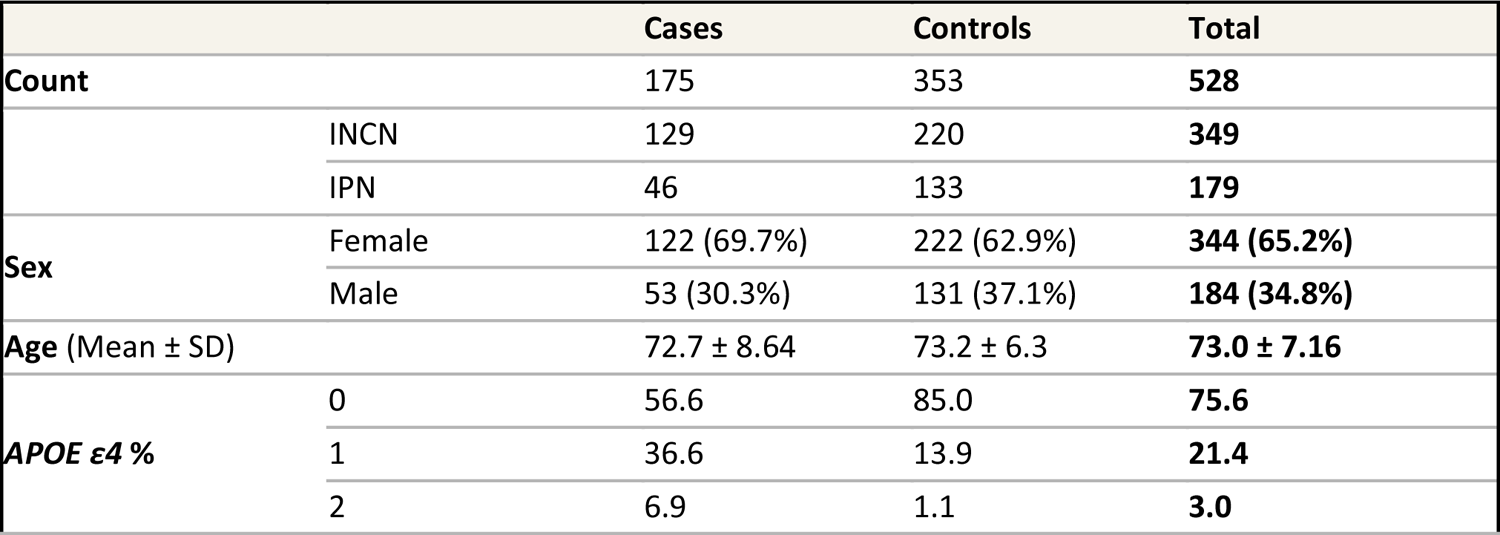
Table showing the age, gender, *APOE ε4* dosage distributions of the participants in our study.

**Table 2.** Results of Single Variant Analysis and PIP Values from Fine-Mapping. Allele frequency (AF) in the table reflects the effect allele frequency in the controls.

|  |  |  |  |  | Model 1 |  |  | Model 2 |  |  |
| --- | --- | --- | --- | --- | --- | --- | --- | --- | --- | --- |
| Closest gene | Marker | dbSNP | Reference /effect allele | AF | OR (95% CI) | P value | PIPs | OR (95% CI) | P value | PIPs |
| Novel Loci |  |  |  |  |  |  |  |  |  |  |
| *NFASC* | 1:204934830 | rs2625222 | T/C | 0.013 | 6.80 (3.39-13.64) | **3.5 × 10<sup>-8</sup>** | 0.39 | 7.64 (3.74-15.61) | **1.2 × 10<sup>-8</sup>** | 0.28 |
| *STK32A* | 5:147301014 | rs4441917 | A/C | 0.422 | 0.45 (0.33-0.61) | 1.3 × 10<sup>-7</sup> | 0.81 | 0.48 (0.35-0.65) | 1.6 × 10<sup>-6</sup> | 0.55 |
| *RP11-663N22.1* | 17:27248016 | rs56281491 | T/G | 0.497 | 2.05 (1.55-2.73) | 3.4 × 10<sup>-7</sup> | 0.83 | 1.97 (1.46-2.64) | 3.7 × 10<sup>-6</sup> | 0.40 |
| AD-known Loci (Bellenguez et al.[5]) |  |  |  |  |  |  |  |  |  |  |
| *APOE* | 19:44908684 | rs429358 | T/C | 0.082 | 3.20 (2.22-4.62) | **2.4 × 10<sup>-10</sup>** | 0.82 | 0.19 (0.01-3.64) | 0.271 | 0 |
| *TREML2* | 6:41181270 | rs60755019 | A/G | 0.041 | 2.12 (1.20-3.75) | 0.005 | 0 | 2.21 (1.22-3.97) | 0.004 | 0 |
| *CLU* | 8:27607795 | rs11787077 | C/T | 0.394 | 0.73 (0.53-1.00) | 0.023 | 0 | 0.74 (0.53-1.03) | 0.039 | 0 |

Single-variant testing replicated the *APOE* locus at P ≤ 5×10^−8^ (Table 2, Fig. 1). In addition, two additional loci known to be associated with AD, *TREML2* (rs60755019, P=0.003) on chromosome 6 and *CLU* (rs11787077, P=0.009) on chromosome 8, showed nominal significance (Table 2). Detailed results for Known-AD variants can be found in the supplemental tables 2 and 3.

As a result of gene-based testing, a locus known to be associated with rare variants in AD, *TREM2* [15], showed nominal significance in the CADD10 and CADD20 variant sets in both the main and *APOE*-adjusted models (S4 Table). There was no gene-wide significant region.

### Pathway analysis

MAGMA gene-set analysis showed no pathway at P_bon_ < .05 after Bonferroni correction (17,009 genes were tested). Three pathways were identified which had P < 1 x 10^-4^, although these pathways were not significant after Bonferroni correction (Table 3).

**Table 3.** Top pathways derived from MAGMA gene-set analysis.

| Pathway name | Number of Genes | P value |  |
| --- | --- | --- | --- |
|  |  | Model 1 | Model 2 |
| Reactome Heparan Sulfate Heparin HS GAG Metabolism | 50 | $8.1 \times 10^{-5}$ | $3.5 \times 10^{-5}$ |
| Reactome Glycosaminoglycan Metabolism | 118 | $1.6 \times 10^{-3}$ | $3.9 \times 10^{-5}$ |
| Reactome Diseases Associated with Glycosaminoglycan Metabolism | 38 | $3.2 \times 10^{-3}$ | $9.7 \times 10^{-5}$ |

### Transcriptomics analyses

Based on the brain-cerebellum data from GTEx v8, the rs2625222 variant, the lead marker in the *NFASC* locus, showed evidence of being an eQTL for *CNTN2* (Contactin-2) with increased expression in variant carriers compared to wildtype carriers (S2 Fig. b). The *CNTN2* gene is located on chromosome 1 approximately 20kb downstream of the *NFASC* gene. In addition, gene expression analysis showed that *APOE*, *NFASC,* and *CNTN2* genes were found to be highly expressed in the brain (S2 Fig. a).

### Functional analysis

rs2625222 is within the second intron of *NFASC*. To understand the potential function of the intronic SNP, we examined chromatin accessibility of the region over rs2625222 in the frontal cortex using snATAC-seq. The variant is located within open ATAC-seq peaks mainly seen in excitatory neurons suggesting that the surrounding DNA sequence is accessible for transcriptional regulation activity (S3 Fig.) [16]. Single nuclei RNAseq data from the frontal cortex demonstrated that NFASC is expressed in oligodendrocytes > neurons [17]. In addition, we examined 3D genome structure using Hi-C analysis in iPSC-derived astrocytes, microglia, oligodendrocytes, and neurons. This analysis revealed cell-specific chromatin loops, which often represent promoter-enhancer interaction. In iPSC-derived neurons and oligodendrocytes but not in iPSC-derived astrocytes and microglial, rs2625222 colocalizes with a major, primary chromatin loop of ∼100kb between the promoter region of *NFASC* and rs2625222 (Figure 2). A minor chromatin loop of ∼380 kb with an intergenic region near the promoter of the long noncoding RNA *ENST0000453895* (Figure 2), which has no reported expression in GTEX in brain. Both loops are absent in astrocytes and microglial. Allele specific Hi-C was not performed. These results show that rs2625222 lies in an active regulatory region in neurons and oligodendrocytes and that the promoter of NFASC is the main target of the associated region with the major Hi-C loop.

**Figure 2.**
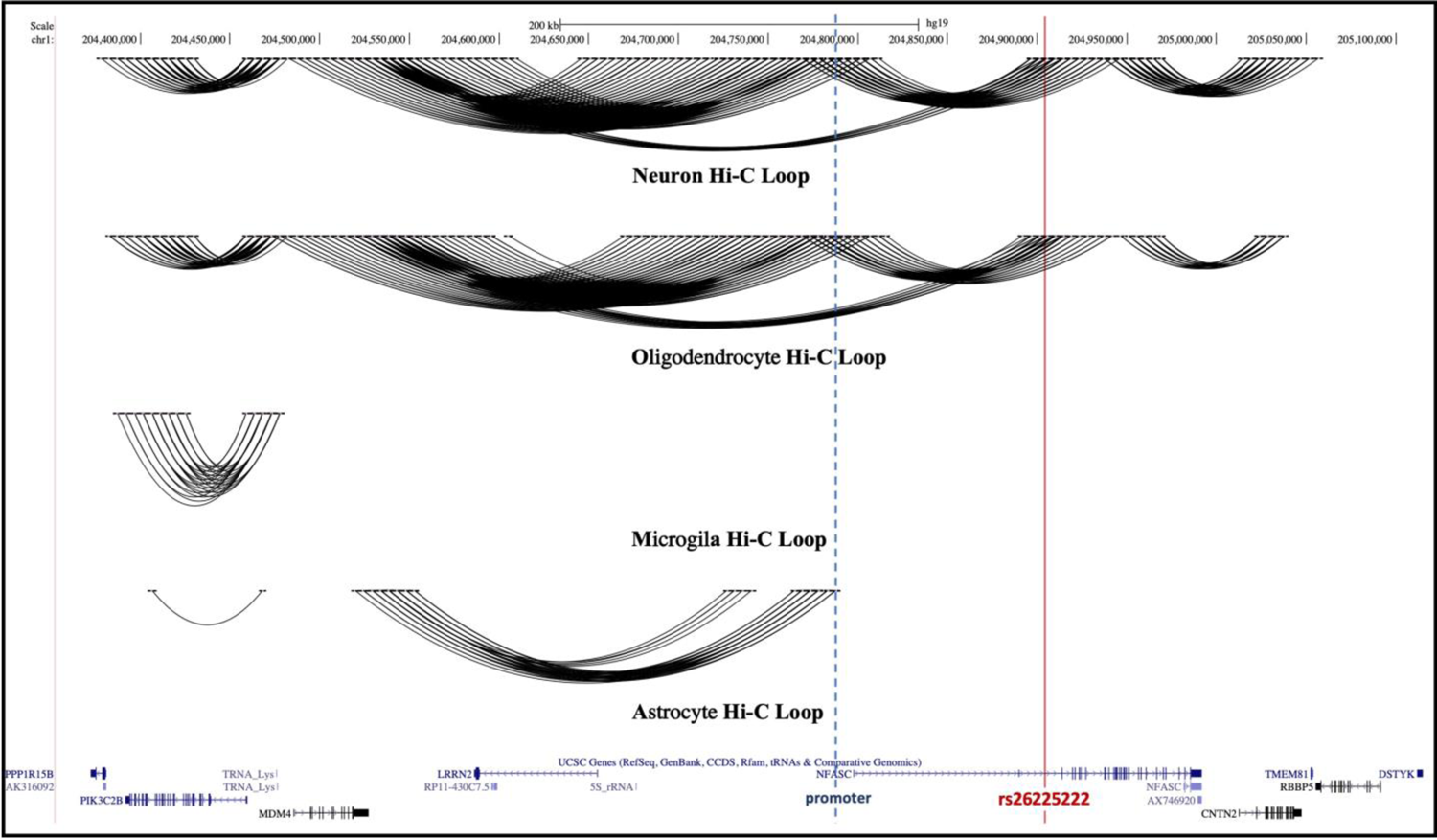
Chromatin Hi-C analysis of the associated region. Interaction loops are observed in Oligodendrocytes and Neurons between the rs26225222 haplotype (vertical red line) and the promoter region of NFASC (dotted line). A minor loop from rs262522 is observed to a region near the promoter of ENST0000453895.

### Fine mapping and ancestry-aware analysis

Seven loci in Models 1 and 2 (three novel risk loci and three AD-known loci) were fine-mapped using CARMA. The corresponding PIPs of each lead SNP are reported in Table 2. A credible set of variants at 0.95 level (i.e., a subset of SNPs that contains a causal variant with the probability of ≥0.95) was generated for the *NFASC* and *APOE* regions. The index SNP within the *NFASC* region (rs2625222) showed the highest PIP value within the region (CADD score = 10.87). On the other hand, there was no credible set generated for *STK32A, RP11-663N22.1,* although these regions’ index SNPs showed higher PIPs compared to that observed for *NFASC*. This effect is likely due to differences in LD patterns between these regions. Specifically, rs2625222 is in high LD with its surrounding SNPs. As a result, the PIP is distributed across these sets of SNPs in high LD, ultimately diluting the PIP for rs2625222; nevertheless, the PIPs’ sum of SNPs in high LD is large enough to formulate a credible set. In contrast, the index SNPs for *STK32A, and RP11-663N22.1* showed weaker LD pattern across adjacent SNPs. Consequently, these SNPs garnered a larger proportion, if not the entirety, of the PIP for their respective regions with the sum of the PIPs of these regions falling short of generating a credible set.

Admixture analysis revealed proportions of 65% AI, 29% EU, 4% AF, and 2% EA in the cohort (S4 Fig.). The local ancestry analysis results of the *NFASC* and *APOE* loci are summarized in Table 4. The ancestry-aware analysis showed the haplotypes associated with AD at the *NFASC* locus were found to be of European origin.

**Table 4.** Local ancestry analysis results of the *NFASC* and *APOE* regions. According to the results of the ancestral dosage extraction of the lead marker in each genome-wide significant region based on 4 populations, the allele frequency within the local ancestry is given in percent, and the allele count is given in parentheses.

|  |  | Alternative Allele Frequency %<br>(Alternative Allele Count) |  |  |  |
| --- | --- | --- | --- | --- | --- |
|  |  | AFR | AI | EAS | EUR |
| **rs2625222**<br>(*NFASC*) | **Controls** | 3.6%<br>(1) | 0%<br>(0) | 0%<br>(0) | 4.2%<br>(8) |
|  | **Cases** | 29.2%<br>(7) | 0%<br>(0) | 0%<br>(0) | 21.1%<br>(23) |
| **rs429358**<br>(*APOE*) | **Controls** | 19.2%<br>(5) | 6.3%<br>(29) | 15.4%<br>(2) | 10.8%<br>(22) |
|  | **Cases** | 45.5%<br>(10) | 17.5%<br>(37) | 33.3%<br>(1) | 34.2%<br>(39) |

Additionally, increased AI ancestral background among controls at the *STK32A* locus was found. This suggests that the *STK32A* locus has a protective effect, particularly among individuals with an AI background (S5 Table). We did not observe significant heterogeneity of effect at the *APOE* and the remaining three novel loci across the different ancestral backgrounds.

### Polygenic risk score

We calculated a PRS using 21,417 clumped SNPs. AUC in PE dataset was found to be 0.63 in Model_PRS-only_ and the t-test showed a significant association between PRS and AD (p = 1.1×10^-5^) (S5 Fig. a). In model*_APOE ε4_*_-only_ and model_PRS +_ *_APOE ε4_*, we achieved an AUC of 0.64 and 0.71 respectively. Model_Full_ showed an AUC of 0.73 (S5 Fig. b).

## Discussion

We identified a novel genome-wide significant signal within the *NFASC gene* on chromosome 1, achieving significance in both Model 1 and Model 2. Our Hi-C, ATAC and expression data strongly support that the *NFASC* gene itself is the primary gene tagged by the risk associated rs2625222 variant. It is certainly possible that rs2625222 associated haplotype may influence other gene promoters, as this is a recognized property of many enhancers. Interestingly, there are strong loops from the promoter region of *NFASC* to the intergenic region near *ENST0000453895*, which may also be involved in regulating this gene. *NFASC* encodes Neurofascin, which plays a crucial role in the development and function of axon initial segments in the central and peripheral nervous systems [18]. Neurofascin is broadly expressed in the brain and is an L1 family immunoglobulin cell adhesion molecule (L1CAM) involved in axon subcellular targeting and synapse formation during neural development [19]. Prior research has shown that increased L1CAM levels in mouse models of AD affect protein kinase D1 activity - which had been found to prevent neuronal cell death-in the cerebral cortex [20]. Pathogenic homozygous missense mutations in the *NFASC* gene have also been associated with a type of neurodevelopmental disorder characterized by central and peripheral motor dysfunction [21].

Ancestry-aware analysis showed that the *NFASC* haplotype conferring increased risk for AD in non-Amerindian (EU and AF) backgrounds. The absence of suggestive or genome-wide significant signals for this locus in previous GWASs for AD could be indicative of a founder effect or gene-environment (GxE) interaction effect in Peru. Evolutionary genomics research [14, 22] on the demographic history of South America and Peru shows recent European and African admixture in this population, which also supports the probability of a young founder mutation in this genetic region. Our functional and ancestry-aware follow-up studies imply that the NFASC region is likely a novel AD susceptibility locus.

Two novel loci reached suggestive significance (P ≤ 5 × 10^−7^), rs4441917 in *STK32A* on chromosome 5 (Model 1) and rs56281491 next to *RP11-663N22.1* on chromosome 17 (Model 1 and 2). The *STK32B* gene, a known paralog of *STK32A*, was associated with the essential tremor [23] and the rate of cognitive decline in AD [24]. Through our ancestry analysis, we observed a significant increase of the rs4441917_C frequency in the control group with AI background whereas we did not observe frequency difference in other ancestral backgrounds, suggesting its protective effects to be AI-ancestry specific.

Our single variant association analysis results replicated same top signals of the three known AD loci[5] – *APOE* (rs429358, *P*=2.4×10^-10^), *TREML2* (rs60755019, *P*=0.005) and *CLU* (rs11787077, *P*=0.023). The *CLU* gene, encoding Clusterin, was first identified as AD risk-gene in 2 independent GWASs [25, 26] and is considered one of the greatest genetic risk factor for AD [27]. Also, a recent and the largest NHW GWAS on AD [5] replicated *TREML2* (rs60755019), and *CLU* (rs11787077) loci. Our local analysis results showed that the signal in the *TREML2* locus was predominantly close to the AI background, and the signal in the *CLU* locus was close to both the AI and EU backgrounds (S5 Table). In the human genome, *TREML2* and closely related *TREM2* gene - an AD risk locus in both NHW [28, 29] and AF [30] populations- are located in the same cluster on chromosome 6. Our gene-based rare variant testing results showed the nominal significance (*P<.05)* of the *TREM2* gene in both models filtered by CADD score of 10 and above and 20 and above, although it did not show the nominal significance of the *TREML2* gene.

NHW GWAS [5]-derived PRS showed a good predictive value (AUC of 0.63 in Model_PRS-only_) of AD risk in the PE population. While the results provide a promising prediction value, there is potential to further optimize the PRS calculations for PE to enhance their clinical relevance. The accuracy of PRS improves when modeled using GWAS with a similar ancestral origin [31]. Therefore, we believe that an AI GWAS-based PRS would likely have improved predictive accuracy compared to PRS derived from NHW GWAS. Nonetheless, it is highly likely that the NHW GWAS-based PRS showed good predictive results is due to the substantial EU ancestral background among Peruvians. Overall, our results point to the importance of performing population-specific studies to derive PRS calculations that will yield high predictive values that are suitable for clinical use.

The poor generalizability of genetic studies across populations is well-established. To understand the myriad genetic factors that contribute to the development of AD it is important to study diverse populations which are underrepresented in genetic studies. By including diverse populations, not only can we identify factors that contribute to health disparities, but we can also fine tune our efforts to develop effective treatments for AD. Further, by including underrepresented populations in genetic studies, higher-sensitivity risks can be calculated with methods such as PRS: more importantly, new genetic loci can be discovered, as in our study, and the biological role of known loci in different populations can be understood more clearly. Thus, a more effective approach to the prevention of AD can be achieved by initiating treatments at the preclinical stage [32], a timing frame when the pathophysiological mechanisms of the disease begin, decades before the clinically detectable symptoms of AD appear [33]. Including underrepresented populations such as the Peruvian population, which has a large proportion of AI background, provides an important opportunity to evaluate the role of AI ancestry in AD, and may pave the way for more accurate prevention, early detection, and intervention of AD in this and other AI predominant admixed populations.

### Limitations

Since our study is the first AD GWAS study on the PE population, the sample size remains lower than in similar NHW-based GWAS studies. Genome-wide studies with larger samples of the AI population are needed to make the effect sizes of this GWAS data more accurate.

## Methods

### Study participants

Participants were ascertained from six different regions in Peru including the Coast (Lima-Callao, Tacna) and Andes highlands (Arequipa, Huancayo-Jauja, Puno, Tacna and Cusco). All ascertainment was coordinated by the Instituto Nacional de Ciencias Neurológicas (INCN), and Instituto Peruano de Neurociencias (IPN).

Informed consent was obtained from all participants, and the study protocols were approved by the Ethical Research Committee of the INCN and IPN Lima, the University of Miami’s Institutional Review Board, and the Columbia University Review Board. Individuals who were deemed eligible at the respective sites were clinically evaluated using standard procedures for genetic studies of AD that were specific to the IPN or INCN ascertainment sites.

All eligible participants underwent an initial screening consisting of a standard clinical interview which included detailed medical and family history as well as the Pfeffer Functional Activities Questionnaire and depending on the ascertainment site, either the Clock Drawing Test (INCN) or the Rowlands Universal Dementia Scale (RUDAS) IPN [34–36]. Individuals who failed the screening were then evaluated with a comprehensive multi-domain cognitive battery which included measures of memory, executive function, language, and visuospatial ability. In addition, these participants were evaluated using functional measures including the Clinical Dementia Rating Scale (CDR). Using all available clinical information, participants were adjudicated by neurologists and neuropsychologists with expertise in neurodegenerative disorders. Clinical research diagnoses were assigned using the National Institute of Aging-Alzheimer’s Association (NIA-AA) criteria for possible and probable AD [37] or the DSM-V criteria for Major Neurocognitive Disorder, Alzheimer’s type [38]. Cases were defined as participants who met NIA-AA or DSM-V criteria for AD. Controls were defined as participants who were cognitively unimpaired and ≥ 65 years of age at study entry.

### Genotyping and quality control

Genome-wide single-nucleotide polymorphism (SNP) genotyping was performed using the Global Screening Array v2.0 (Illumina, San Diego, CA, USA). *APOE* genotyping was performed as in Saunders et al [39].

For quality control, genotyped data were cleaned with standard QC measures using PLINK software [40]. In brief, SNPs with Hardy–Weinberg equilibrium p-value>1×10^-6^ and genotype missingness ≥ 3% were excluded, and individuals with genotype missingness ≥ 3% were also excluded. To ensure the accuracy and reliability of the results, only variants with minor allele count (MAC) ≥ 5 were included in the analysis. Principal components (PCs) were calculated using the Eigenstrat program [41]. To determine the PCs used for further analyses, we employed logistic regression modelling (AD ∼ Sex + Age + PC1:10).

### Genotype imputation

After genotype quality control, cleaned genotyped data was imputed to NHLBI TOPMed Freeze 5 haplotype reference panel [42], which includes around 400M variants observed from 53,831 individuals, using the TOPMed Imputation Server. All reference population haplotypes were used for the imputation. Common variants (MAF ≥ 0.01) with an R^2^< 0.40 and rare variants (MAF < 0.01) with an R^2^ <0.80 were excluded from further analyses.

### Association analysis

#### Single variant analysis

Single variant association analysis was performed using SAIGE [43] on genotype dosages employing a linear mixed model. We analyzed the data in two separate models; the first model accounted for sex, age, and PCs for population substructure (Model 1), while the second model also included the dosage of the *APOE ε4* allele (Model 2). In both models, we included a genetic relationship matrix as a random effect to account for any potential relatedness. The GenABEL package version 1.8-031 was used to estimate genomic inflation (λ).

#### Known AD genes

Known AD loci were determined from the latest AF[30] and NHW[5] GWASs. Same top signals were extracted from our association analysis results for both models and evaluated whether they were replicated or not based on the P value threshold of .05.

#### Gene-based analysis

Before the gene-based test, variants were restricted to rare variants excluding all variants with minor allele frequency (MAF) > 0.05 and R^2^ < 0.8. Then, variants were annotated with AnnoVar[44] to identify the gene region and the CADD [45] score. As a result of gene region annotation, only intragenic variants (upstream, downstream, exonic and intronic variants) were included in the analysis. A combined test of burden and sequence kernel association test (SKAT-O) [46] was performed using the GMMAT tool [47]. Three different variant sets were assessed: CADD20 set (variants with a CADD score of 20 or higher), CADD10 set (variants with a CADD score of 10 or higher) and CADD0 set (all intragenic variants). All sets were tested twice with two models: a main model (adjusted for sex, age, and first 4 PCs as fixed effects and GRM as a random effect), and an additional *APOE ε4* allele dosage adjusted model.

### Pathway analysis

We performed pathway analyses with Multi-marker Analysis of GenoMic Annotation (MAGMA) [48] v1.08 using FUMA [49] v1.5.6, which performs SNP-wise gene analysis. 15496 gene sets (Curated gene sets: 5500, Gene Ontology (GO) terms: 9996) obtained from MsigDB [50, 51] v7.0 were used in the analyses. We analyzed a 35-kb upstream and 10-kb downstream window around each gene.

### Transcriptomics analysis

Expression quantitative trait loci (eQTL) mapping, which maps SNPs that likely affect the expression of a target gene(s) and lies up to 1 Mb (cis-eQTL) away from the target gene, was performed using FUMA. GTEx v8 [52] Brain Tissue Types was used for eQTL mapping.

### Functional analysis

#### Hi-C chromatin studies

Hi-C analysis data (unpublished) was obtained on iPSC-derived astrocytes, microglial, oligodendrocyte-enriched spheroids, and neurons created as part of ongoing studies in Alzheimer disease (S1 Method).

In situ Hi-C libraries were created based on the protocol from Rao et al [53]. In brief, for each library, 450∼550 million paired-end reads of 150 bp length were obtained. Over 270 million non-redundant, uniquely mapped, paired reads were used for further analysis of each library. Contact matrices were generated at base pair delimited resolutions of 500 kb, 50 kb, and 5 kb. The Directionality Index (DI) value of 40kb bins was used to call the topological associated domain (TAD). For robust enhancer-promoter interaction mapping, the HiCorr pipeline was used to correct Hi-C bias at sub-TAD level to identify Hi-C loops [54]. Detailed methods for iPSC, cell differentiation and Hi-C can be found in the supplemental methods.

#### RNA expression

Data was obtained from previously published single nuclei ATACseq (snATACseq) and single nuclei RNAseq (snRNAseq) studies of the frontal cortex (B9) from African and European ApoE4 carriers [16, 17].

### Fine-mapping and ancestral aware analysis

#### Fine-mapping

Fine-mapping was performed using CARMA [55] with each locus defined as a 1Mb region centered around the index SNP with suggestive significance (P<5×10^-7^) and AD known loci [5]. Each locus’ LD matrix was generated based on the individual-level genetic data used in the association analysis. We employed CARMA with default values for all parameters with the maximum number of causal variants assumed in a region set at N=10. The functional annotation CADD [45] and SEI [56], which learn a vocabulary of regulatory activities through a deep learning approach, were also provided to CARMA as prior information on the causality of the testing SNPs. The credible sets generated by CARMA were reported in the (S1 Table) with the corresponding posterior inclusion probability (PIP) that indicates the causality of SNP considering the local LD structure and CADD and SEI scores.

#### Global ancestry estimation

The admixture proportion was estimated by using a model-based clustering algorithm implemented in the ADMIXTURE software [57]. Supervised ADMIXTURE analysis was performed at K = 4 by including the 4 reference populations (AI, EU, AF, and EA) from combined reference panels of the Human Genome Diversity Project (HGDP) [58] and 1000 Genomes Phase 3 [59, 60].

#### Local ancestry estimation

The local ancestry was assessed by combining the 4 populations (AI, EU, AF, and EA) in combined reference panels of HGDP [58] and 1000 Genomes Phase 3 [59, 60] with the imputed Peruvian dataset. The SHAPEIT [61] tool was used to phase all individuals in the same combined reference panels, and the RFMix Version 2 [62] tool with the discriminative modelling approach was used to infer the local ancestry at each locus across the genome. The standard parameters were used with a minimum node size of 5 in order to perform RFMix analysis.

#### Local ancestry analysis

After the local ancestry estimation step, ancestry dosages for variants of interest based on 4 population was extracted using Tractor [63] for lead markers in each loci that were both genome-wide significant and suggestive according to the association analysis of each individual. These individuals were divided into four groups according to their ancestral background for each allele at each locus of interest: AF, AI, EA, and EU. Then, allele frequencies and allele count of cases and controls were calculated for each subgroup according to the lead markers at the loci (Table 4).

### Polygenic risk score

We constructed PRS on the PE dataset using the effect sizes from summary statistics from the largest NHW GWAS study [5]. Quality control steps were carried out using standard parameters in the literature [31]. We removed duplicate and ambiguous SNPs from the summary statistics NHW GWAS with the custom script. For the PE dataset, SNPs were excluded if MAF ≤0.01%, Hardy–Weinberg equilibrium p <1×10^-6^, and genotype missingness ≥1%; individuals were removed if genotype missingness ≥1% using PLINK (v1.9) [40].

The PRSice-2 [64] tool was used to generate the PRS. Analyses were performed with standard parameters in accordance with the published PRS tutorial [31]. We applied LD-clumping using the following parameters: -- clump-kb 250 – clump-r2 0.1 –clump p1. We also filtered out variants with minor allele frequency (MAF) was less than 5%. We included only autosomal chromosomes in the analysis. In order evaluate PRS performance independent of *APOE* effect, we first removed the *APOE* region (2 MB around *APOE ε4* SNP) from the data. Then, to adjust the model, we used age, sex, and the first three PCs as covariates.

After each PRS calculation, the PRS performance was assessed by employing the logistic regression model: PRS-only, *APOE ε4*-only, PRS + *APOE ε4*, and Full (PRS+ *APOE ε4* + Sex + Age + PC1:3) to construct receiver operator curves (ROC).

1. AD ∼ PRS (“Model_PRS-only_”)
2. AD ∼ *APOE ε4* (“Model*_APOE ε4_*_-only_”)
3. AD ∼ PRS + *APOE ε4* (“Model_PRS +_ *_APOE ε4_*,”)
4. AD ∼ PRS + *APOE ε4* + Sex + Age + PC1:3 (“Model_Full_”)

## Supporting information

Supplemental Content

## Data Availability

Data are available through the National Institute on Aging Genetics of Alzheimers Disease Data Storage Site (NIAGADS) Data Sharing Service (DSS).

https://dss.niagads.org/datasets/ng00067/

## Acknowledgements

We are grateful the DNA-Neurogenetics Bank of the Instituto Nacional de Ciencias Neurológicas for supporting sample and data management for INCN-based participants, Neurogenetics Research Unit at San Marcos Foundation for administrative and logistic support, and all participants and their families for taking part in our study.

## Supporting information

**S1 Method.** Functional analysis

**S1 Fig.** Quantile-quantile plots show the deviation observed from expected p-values of single variant association analysis.

**S2 Fig.** a) Gene expression heatmap of eQTL mapped genes at P ≤ 1 × 10^-6^. b) Single tissue (Brain - Cerebellar Hemisphere) eQTL violin plot of rs2625222 for *CNTN2* expression.

**S3 Fig.** Chromatin accessibility analysis of the region surrounding rs2625222 in frontal cortex. Visualization of chromatin accessibility by ATAC-seq peaks in the frontal cortex.6 rs2625222 is indicated as the red vertical line.

**S4 Fig.** The global ancestry bar plot of the 4-way admixed Peruvian individuals in our study.

**S5 Fig.** a) Boxplots showing the relationship of NHW GWAS-based PRS results between the case and control group. b) Figure showing ROC curves of logistic regression models of different models.

**S6 Fig.** Regional association plots, generated using LocusZoom.js[65], for the two genome-wide significant and two suggestive loci based on Model 1: **a)** *NFASC* on chromosome 1, **b)** *STK32A* on chromosome 5, **c)** *RP11-663N22.1* on chromosome 17, and **d)** *APOE* on chromosome 19. Each regional plot labels the SNPs with the lowest P value at each locus, depicting them as purple diamonds. The triangles represent individual SNPs, and the colors of the triangles indicate LD with the SNP with the lowest P value, based on the AMR reference population. The right-hand y-axis of each regional plot displays the recombination rate, marked by blue vertical lines.

**S1 Table.** The credible sets generated by CARMA.

**S2 Table.** Single-variant testing results for Known AD variants in Model 1.

**S3 Table.** Single-variant testing results for Known AD variants in Model 2.

**S4 Table.** Nominal significant genes as a result of rare variant gene-based testing for Model 1 (adjusted for sex, age, and first 4 PCs as fixed effects and GRM as a random effect), and Model 2 (also adjusted for APOE-ε4 allele dosage).

**S5 Table.** Local ancestry analysis results of the *NFASC*, *APOE*, two novel suggestive, and replicated Known-AD loci. According to the results of the ancestral dosage extraction of the lead marker in each genome-wide significant region based on 4 populations, the allele frequency is given in percent, and the allele count is given in parentheses.

## References

1. 2020 Alzheimer’s disease facts and figures. Alzheimers Dement. 2020. Epub 20200310. doi: 10.1002/alz.12068. PubMed PMID: 32157811.

2. Borenstein AR, Mortimer JA, ScienceDirect. Alzheimer’s disease: life course perspectives on risk reduction. Amsterdam: Academic Press is an imprint of Elsevier; 2016.

3. Gatz M, Pedersen NL, Berg S, Johansson B, Johansson K, Mortimer JA, et al. Heritability for Alzheimer’s disease: the study of dementia in Swedish twins. J Gerontol A Biol Sci Med Sci. 1997;52(2):M117–25. doi: 10.1093/gerona/52a.2.m117. PubMed PMID: 9060980.

4. Gatz M, Reynolds CA, Fratiglioni L, Johansson B, Mortimer JA, Berg S, et al. Role of genes and environments for explaining Alzheimer disease. Arch Gen Psychiatry. 2006;63(2):168–74. doi: 10.1001/archpsyc.63.2.168. PubMed PMID: 16461860.

5. Bellenguez C, Kucukali F, Jansen IE, Kleineidam L, Moreno-Grau S, Amin N, et al. New insights into the genetic etiology of Alzheimer’s disease and related dementias. Nat Genet. 2022;54(4):412–36. Epub 20220404. doi: 10.1038/s41588-022-01024-z. PubMed PMID: 35379992; PubMed Central PMCID: PMCPMC9005347.

6. Mills MC, Rahal C. A scientometric review of genome-wide association studies. Commun Biol. 2019;2:9. Epub 20190107. doi: 10.1038/s42003-018-0261-x. PubMed PMID: 30623105; PubMed Central PMCID: PMCPMC6323052.

7. Mills MC, Rahal C. The GWAS Diversity Monitor tracks diversity by disease in real time. Nat Genet. 2020;52(3):242–3. doi: 10.1038/s41588-020-0580-y. PubMed PMID: 32139905.

8. Cukier HN, Kunkle BW, Vardarajan BN, Rolati S, Hamilton-Nelson KL, Kohli MA, et al. ABCA7 frameshift deletion associated with Alzheimer disease in African Americans. Neurol Genet. 2016;2(3):e79. Epub 20160517. doi: 10.1212/NXG.0000000000000079. PubMed PMID: 27231719; PubMed Central PMCID: PMCPMC4871806.

9. Farrer LA, Cupples LA, Haines JL, Hyman B, Kukull WA, Mayeux R, et al. Effects of age, sex, and ethnicity on the association between apolipoprotein E genotype and Alzheimer disease. A meta-analysis. APOE and Alzheimer Disease Meta Analysis Consortium. JAMA. 1997;278(16):1349–56. PubMed PMID: 9343467.

10. Liu F, Ikram MA, Janssens AC, Schuur M, de Koning I, Isaacs A, et al. A study of the SORL1 gene in Alzheimer’s disease and cognitive function. J Alzheimers Dis. 2009;18(1):51–64. doi: 10.3233/JAD-2009-1137. PubMed PMID: 19584446.

11. Reitz C, Jun G, Naj A, Rajbhandary R, Vardarajan BN, Wang LS, et al. Variants in the ATP-binding cassette transporter (ABCA7), apolipoprotein E ɛ4,and the risk of late-onset Alzheimer disease in African Americans. JAMA. 2013;309(14):1483–92. doi: 10.1001/jama.2013.2973. PubMed PMID: 23571587; PubMed Central PMCID: PMCPMC3667653.

12. Reitz C, Pericak-Vance MA, Foroud T, Mayeux R. A global view of the genetic basis of Alzheimer disease. Nat Rev Neurol. 2023. Epub 20230406. doi: 10.1038/s41582-023-00789-z. PubMed PMID: 37024647.

13. Custodio N, García A, Montesinos R, Escobar J, Bendezú L. Prevalencia de demencia en una población urbana de Lima-Perú: estudio puerta a puerta. An Fac med. 2008;69(4):233–8. doi: 10.15381/anales.v69i4.1110.

14. Harris DN, Song W, Shetty AC, Levano KS, Caceres O, Padilla C, et al. Evolutionary genomic dynamics of Peruvians before, during, and after the Inca Empire. Proc Natl Acad Sci U S A. 2018;115(28):E6526-E35. Epub 20180626. doi: 10.1073/pnas.1720798115. PubMed PMID: 29946025; PubMed Central PMCID: PMCPMC6048481.

15. Sims R, van der Lee SJ, Naj AC, Bellenguez C, Badarinarayan N, Jakobsdottir J, et al. Rare coding variants in PLCG2, ABI3, and TREM2 implicate microglial-mediated innate immunity in Alzheimer’s disease. Nat Genet. 2017;49(9):1373–84. Epub 20170717. doi: 10.1038/ng.3916. PubMed PMID: 28714976; PubMed Central PMCID: PMCPMC5669039.

16. Celis K, Moreno M, Rajabli F, Whitehead P, Hamilton-Nelson K, Dykxhoorn DM, et al. Ancestry-related differences in chromatin accessibility and gene expression of APOE epsilon4 are associated with Alzheimer’s disease risk. Alzheimers Dement. 2023. Epub 20230410. doi: 10.1002/alz.13075. PubMed PMID: 37037656.

17. Griswold AJ, Celis K, Bussies PL, Rajabli F, Whitehead PL, Hamilton-Nelson KL, et al. Increased APOE epsilon4 expression is associated with the difference in Alzheimer’s disease risk from diverse ancestral backgrounds. Alzheimers Dement. 2021;17(7):1179–88. Epub 20210201. doi: 10.1002/alz.12287. PubMed PMID: 33522086; PubMed Central PMCID: PMCPMC8843031.

18. Smigiel R, Sherman DL, Rydzanicz M, Walczak A, Mikolajkow D, Krolak-Olejnik B, et al. Homozygous mutation in the Neurofascin gene affecting the glial isoform of Neurofascin causes severe neurodevelopment disorder with hypotonia, amimia and areflexia. Hum Mol Genet. 2018;27(21):3669–74. doi: 10.1093/hmg/ddy277. PubMed PMID: 30124836; PubMed Central PMCID: PMCPMC6196652.

19. Ango F, di Cristo G, Higashiyama H, Bennett V, Wu P, Huang ZJ. Ankyrin-based subcellular gradient of neurofascin, an immunoglobulin family protein, directs GABAergic innervation at purkinje axon initial segment. Cell. 2004;119(2):257–72. doi: 10.1016/j.cell.2004.10.004. PubMed PMID: 15479642.

20. Chen S, Jiang Q, Huang P, Hu C, Shen H, Schachner M, et al. The L1 cell adhesion molecule affects protein kinase D1 activity in the cerebral cortex in a mouse model of Alzheimer’s disease. Brain Res Bull. 2020;162:141–50. Epub 20200612. doi: 10.1016/j.brainresbull.2020.06.004. PubMed PMID: 32540419.

21. Anazi S, Maddirevula S, Salpietro V, Asi YT, Alsahli S, Alhashem A, et al. Expanding the genetic heterogeneity of intellectual disability. Hum Genet. 2017;136(11-12):1419–29. Epub 20170922. doi: 10.1007/s00439-017-1843-2. PubMed PMID: 28940097.

22. Homburger JR, Moreno-Estrada A, Gignoux CR, Nelson D, Sanchez E, Ortiz-Tello P, et al. Genomic Insights into the Ancestry and Demographic History of South America. PLoS Genet. 2015;11(12):e1005602. Epub 20151204. doi: 10.1371/journal.pgen.1005602. PubMed PMID: 26636962; PubMed Central PMCID: PMCPMC4670080.

23. Liao C, Sarayloo F, Vuokila V, Rochefort D, Akcimen F, Diamond S, et al. Transcriptomic Changes Resulting From STK32B Overexpression Identify Pathways Potentially Relevant to Essential Tremor. Front Genet. 2020;11:813. Epub 20200731. doi: 10.3389/fgene.2020.00813. PubMed PMID: 32849812; PubMed Central PMCID: PMCPMC7413243.

24. Sherva R, Tripodis Y, Bennett DA, Chibnik LB, Crane PK, de Jager PL, et al. Genome-wide association study of the rate of cognitive decline in Alzheimer’s disease. Alzheimers Dement. 2014;10(1):45–52. Epub 20130325. doi: 10.1016/j.jalz.2013.01.008. PubMed PMID: 23535033; PubMed Central PMCID: PMCPMC3760995.

25. Harold D, Abraham R, Hollingworth P, Sims R, Gerrish A, Hamshere ML, et al. Genome-wide association study identifies variants at CLU and PICALM associated with Alzheimer’s disease. Nat Genet. 2009;41(10):1088–93. Epub 20090906. doi: 10.1038/ng.440. PubMed PMID: 19734902; PubMed Central PMCID: PMCPMC2845877.

26. Lambert JC, Heath S, Even G, Campion D, Sleegers K, Hiltunen M, et al. Genome-wide association study identifies variants at CLU and CR1 associated with Alzheimer’s disease. Nat Genet. 2009;41(10):1094–9. Epub 20090906. doi: 10.1038/ng.439. PubMed PMID: 19734903.

27. Foster EM, Dangla-Valls A, Lovestone S, Ribe EM, Buckley NJ. Clusterin in Alzheimer’s Disease: Mechanisms, Genetics, and Lessons From Other Pathologies. Front Neurosci. 2019;13:164. Epub 20190228. doi: 10.3389/fnins.2019.00164. PubMed PMID: 30872998; PubMed Central PMCID: PMCPMC6403191.

28. Guerreiro R, Wojtas A, Bras J, Carrasquillo M, Rogaeva E, Majounie E, et al. TREM2 variants in Alzheimer’s disease. N Engl J Med. 2013;368(2):117–27. Epub 20121114. doi: 10.1056/NEJMoa1211851. PubMed PMID: 23150934; PubMed Central PMCID: PMCPMC3631573.

29. Jonsson T, Stefansson H, Steinberg S, Jonsdottir I, Jonsson PV, Snaedal J, et al. Variant of TREM2 associated with the risk of Alzheimer’s disease. N Engl J Med. 2013;368(2):107–16. Epub 20121114. doi: 10.1056/NEJMoa1211103. PubMed PMID: 23150908; PubMed Central PMCID: PMCPMC3677583.

30. Kunkle BW, Schmidt M, Klein HU, Naj AC, Hamilton-Nelson KL, Larson EB, et al. Novel Alzheimer Disease Risk Loci and Pathways in African American Individuals Using the African Genome Resources Panel: A Meta-analysis. JAMA Neurol. 2021;78(1):102–13. doi: 10.1001/jamaneurol.2020.3536. PubMed PMID: 33074286; PubMed Central PMCID: PMCPMC7573798.

31. Choi SW, Mak TS, O’Reilly PF. Tutorial: a guide to performing polygenic risk score analyses. Nat Protoc. 2020;15(9):2759–72. Epub 20200724. doi: 10.1038/s41596-020-0353-1. PubMed PMID: 32709988; PubMed Central PMCID: PMCPMC7612115.

32. Andrieu S, Coley N, Lovestone S, Aisen PS, Vellas B. Prevention of sporadic Alzheimer’s disease: lessons learned from clinical trials and future directions. Lancet Neurol. 2015;14(9):926–44. Epub 20150723. doi: 10.1016/S1474-4422(15)00153-2. PubMed PMID: 26213339.

33. Sperling R, Mormino E, Johnson K. The evolution of preclinical Alzheimer’s disease: implications for prevention trials. Neuron. 2014;84(3):608–22. Epub 20141105. doi: 10.1016/j.neuron.2014.10.038. PubMed PMID: 25442939; PubMed Central PMCID: PMCPMC4285623.

34. Manos PJ, Wu R. The ten point clock test: a quick screen and grading method for cognitive impairment in medical and surgical patients. Int J Psychiatry Med. 1994;24(3):229–44. doi: 10.2190/5A0F-936P-VG8N-0F5R. PubMed PMID: 7890481.

35. Pfeffer RI, Kurosaki TT, Harrah CH, Jr., Chance JM, Filos S. Measurement of functional activities in older adults in the community. J Gerontol. 1982;37(3):323–9. doi: 10.1093/geronj/37.3.323. PubMed PMID: 7069156.

36. Storey JE, Rowland JT, Basic D, Conforti DA, Dickson HG. The Rowland Universal Dementia Assessment Scale (RUDAS): a multicultural cognitive assessment scale. Int Psychogeriatr. 2004;16(1):13–31. doi: 10.1017/s1041610204000043. PubMed PMID: 15190994.

37. McKhann GM, Knopman DS, Chertkow H, Hyman BT, Jack CR, Jr., Kawas CH, et al. The diagnosis of dementia due to Alzheimer’s disease: recommendations from the National Institute on Aging-Alzheimer’s Association workgroups on diagnostic guidelines for Alzheimer’s disease. Alzheimers Dement. 2011;7(3):263–9. Epub 20110421. doi: 10.1016/j.jalz.2011.03.005. PubMed PMID: 21514250; PubMed Central PMCID: PMCPMC3312024.

38. Association AP. Diagnostic and statistical manual of mental disorders: DSM-5. 5th ed. ed. Arlington, VA:: American Psychiatric Association; 2013.

39. Saunders AM, Hulette O, Welsh-Bohmer KA, Schmechel DE, Crain B, Burke JR, et al. Specificity, sensitivity, and predictive value of apolipoprotein-E genotyping for sporadic Alzheimer’s disease. Lancet. 1996;348(9020):90-3. doi: 10.1016/s0140-6736(96)01251-2. PubMed PMID: 8676723.

40. Purcell S, Neale B, Todd-Brown K, Thomas L, Ferreira MA, Bender D, et al. PLINK: a tool set for whole-genome association and population-based linkage analyses. Am J Hum Genet. 2007;81(3):559–75. Epub 20070725. doi: 10.1086/519795. PubMed PMID: 17701901; PubMed Central PMCID: PMCPMC1950838.

41. Price AL, Patterson NJ, Plenge RM, Weinblatt ME, Shadick NA, Reich D. Principal components analysis corrects for stratification in genome-wide association studies. Nat Genet. 2006;38(8):904–9. Epub 20060723. doi: 10.1038/ng1847. PubMed PMID: 16862161.

42. Taliun D, Harris DN, Kessler MD, Carlson J, Szpiech ZA, Torres R, et al. Sequencing of 53,831 diverse genomes from the NHLBI TOPMed Program. Nature. 2021;590(7845):290-9. Epub 20210210. doi: 10.1038/s41586-021-03205-y. PubMed PMID: 33568819; PubMed Central PMCID: PMCPMC7875770.

43. Zhou W, Zhao Z, Nielsen JB, Fritsche LG, LeFaive J, Gagliano Taliun SA, et al. Scalable generalized linear mixed model for region-based association tests in large biobanks and cohorts. Nat Genet. 2020;52(6):634–9. Epub 20200518. doi: 10.1038/s41588-020-0621-6. PubMed PMID: 32424355; PubMed Central PMCID: PMCPMC7871731.

44. Wang K, Li M, Hakonarson H. ANNOVAR: functional annotation of genetic variants from high-throughput sequencing data. Nucleic Acids Res. 2010;38(16):e164. Epub 20100703. doi: 10.1093/nar/gkq603. PubMed PMID: 20601685; PubMed Central PMCID: PMCPMC2938201.

45. Kircher M, Witten DM, Jain P, O’Roak BJ, Cooper GM, Shendure J. A general framework for estimating the relative pathogenicity of human genetic variants. Nat Genet. 2014;46(3):310–5. Epub 20140202. doi: 10.1038/ng.2892. PubMed PMID: 24487276; PubMed Central PMCID: PMCPMC3992975.

46. Lee S, Emond MJ, Bamshad MJ, Barnes KC, Rieder MJ, Nickerson DA, et al. Optimal unified approach for rare-variant association testing with application to small-sample case-control whole-exome sequencing studies. Am J Hum Genet. 2012;91(2):224–37. Epub 20120802. doi: 10.1016/j.ajhg.2012.06.007. PubMed PMID: 22863193; PubMed Central PMCID: PMCPMC3415556.

47. Chen H, Huffman JE, Brody JA, Wang C, Lee S, Li Z, et al. Efficient Variant Set Mixed Model Association Tests for Continuous and Binary Traits in Large-Scale Whole-Genome Sequencing Studies. Am J Hum Genet. 2019;104(2):260–74. Epub 20190110. doi: 10.1016/j.ajhg.2018.12.012. PubMed PMID: 30639324; PubMed Central PMCID: PMCPMC6372261.

48. de Leeuw CA, Mooij JM, Heskes T, Posthuma D. MAGMA: generalized gene-set analysis of GWAS data. PLoS Comput Biol. 2015;11(4):e1004219. Epub 20150417. doi: 10.1371/journal.pcbi.1004219. PubMed PMID: 25885710; PubMed Central PMCID: PMCPMC4401657.

49. Watanabe K, Taskesen E, van Bochoven A, Posthuma D. Functional mapping and annotation of genetic associations with FUMA. Nat Commun. 2017;8(1):1826. Epub 20171128. doi: 10.1038/s41467-017-01261-5. PubMed PMID: 29184056; PubMed Central PMCID: PMCPMC5705698.

50. Liberzon A, Birger C, Thorvaldsdottir H, Ghandi M, Mesirov JP, Tamayo P. The Molecular Signatures Database (MSigDB) hallmark gene set collection. Cell Syst. 2015;1(6):417–25. doi: 10.1016/j.cels.2015.12.004. PubMed PMID: 26771021; PubMed Central PMCID: PMCPMC4707969.

51. Subramanian A, Tamayo P, Mootha VK, Mukherjee S, Ebert BL, Gillette MA, et al. Gene set enrichment analysis: a knowledge-based approach for interpreting genome-wide expression profiles. Proc Natl Acad Sci U S A. 2005;102(43):15545–50. Epub 20050930. doi: 10.1073/pnas.0506580102. PubMed PMID: 16199517; PubMed Central PMCID: PMCPMC1239896.

52. Carithers LJ, Ardlie K, Barcus M, Branton PA, Britton A, Buia SA, et al. A Novel Approach to High-Quality Postmortem Tissue Procurement: The GTEx Project. Biopreserv Biobank. 2015;13(5):311–9. doi: 10.1089/bio.2015.0032. PubMed PMID: 26484571; PubMed Central PMCID: PMCPMC4675181.

53. Rao SS, Huntley MH, Durand NC, Stamenova EK, Bochkov ID, Robinson JT, et al. A 3D map of the human genome at kilobase resolution reveals principles of chromatin looping. Cell. 2014;159(7):1665–80. Epub 20141211. doi: 10.1016/j.cell.2014.11.021. PubMed PMID: 25497547; PubMed Central PMCID: PMCPMC5635824.

54. Lu L, Liu X, Huang WK, Giusti-Rodriguez P, Cui J, Zhang S, et al. Robust Hi-C Maps of Enhancer-Promoter Interactions Reveal the Function of Non-coding Genome in Neural Development and Diseases. Mol Cell. 2020;79(3):521–34 e15. Epub 20200626. doi: 10.1016/j.molcel.2020.06.007. PubMed PMID: 32592681; PubMed Central PMCID: PMCPMC7415676.

55. Yang Z, Wang C, Liu L, Khan A, Lee A, Vardarajan B, et al. CARMA is a new Bayesian model for fine-mapping in genome-wide association meta-analyses. Nat Genet. 2023;55(6):1057–65. Epub 20230511. doi: 10.1038/s41588-023-01392-0. PubMed PMID: 37169873.

56. Chen KM, Wong AK, Troyanskaya OG, Zhou J. A sequence-based global map of regulatory activity for deciphering human genetics. Nat Genet. 2022;54(7):940–9. Epub 20220711. doi: 10.1038/s41588-022-01102-2. PubMed PMID: 35817977; PubMed Central PMCID: PMCPMC9279145.

57. Zhou H, Alexander D, Lange K. A quasi-Newton acceleration for high-dimensional optimization algorithms. Stat Comput. 2011;21(2):261–73. doi: 10.1007/s11222-009-9166-3. PubMed PMID: 21359052; PubMed Central PMCID: PMCPMC3045213.

58. Fairley S, Lowy-Gallego E, Perry E, Flicek P. The International Genome Sample Resource (IGSR) collection of open human genomic variation resources. Nucleic Acids Res. 2020;48(D1):D941–D7. doi: 10.1093/nar/gkz836. PubMed PMID: 31584097; PubMed Central PMCID: PMCPMC6943028.

59. Delaneau O, Marchini J, Genomes Project C, Genomes Project C. Integrating sequence and array data to create an improved 1000 Genomes Project haplotype reference panel. Nat Commun. 2014;5:3934. Epub 20140613. doi: 10.1038/ncomms4934. PubMed PMID: 25653097; PubMed Central PMCID: PMCPMC4338501.

60. Genomes Project C, Auton A, Brooks LD, Durbin RM, Garrison EP, Kang HM, et al. A global reference for human genetic variation. Nature. 2015;526(7571):68-74. doi: 10.1038/nature15393. PubMed PMID: 26432245; PubMed Central PMCID: PMCPMC4750478.

61. Delaneau O, Marchini J, Zagury JF. A linear complexity phasing method for thousands of genomes. Nat Methods. 2011;9(2):179–81. Epub 20111204. doi: 10.1038/nmeth.1785. PubMed PMID: 22138821.

62. Maples BK, Gravel S, Kenny EE, Bustamante CD. RFMix: a discriminative modeling approach for rapid and robust local-ancestry inference. Am J Hum Genet. 2013;93(2):278–88. Epub 20130801. doi: 10.1016/j.ajhg.2013.06.020. PubMed PMID: 23910464; PubMed Central PMCID: PMCPMC3738819.

63. Atkinson EG, Maihofer AX, Kanai M, Martin AR, Karczewski KJ, Santoro ML, et al. Tractor uses local ancestry to enable the inclusion of admixed individuals in GWAS and to boost power. Nat Genet. 2021;53(2):195–204. Epub 20210118. doi: 10.1038/s41588-020-00766-y. PubMed PMID: 33462486; PubMed Central PMCID: PMCPMC7867648.

64. Choi SW, O’Reilly PF. PRSice-2: Polygenic Risk Score software for biobank-scale data. Gigascience. 2019;8(7). doi: 10.1093/gigascience/giz082. PubMed PMID: 31307061; PubMed Central PMCID: PMCPMC6629542.

65. Boughton AP, Welch RP, Flickinger M, VandeHaar P, Taliun D, Abecasis GR, et al. LocusZoom.js: interactive and embeddable visualization of genetic association study results. Bioinformatics. 2021;37(18):3017–8. doi: 10.1093/bioinformatics/btab186. PubMed PMID: 33734315; PubMed Central PMCID: PMCPMC8479674.

