## Supplemental Content for "A genome-wide association study in Peruvians suggests new risk loci for Alzheimer Disease"

**S1 Method.** Functional analysis

**S1 Fig.** Quantile-quantile plots show the deviation observed from expected p-values of single variant association analysis.

**S2 Fig. a)** Gene expression heatmap of eQTL mapped genes at $P \leq 1 \times 10^{-6}$. **b)** Single tissue (Brain - Cerebellar Hemisphere) eQTL violin plot of rs2625222 for *CNTN2* expression.

**S3 Fig.** Chromatin accessibility analysis of the region surrounding rs2625222 in frontal cortex. Visualization of chromatin accessibility by ATAC-seq peaks in the frontal cortex.6 rs2625222 is indicated as the red vertical line.

**S4 Fig.** The global ancestry bar plot of the 4-way admixed Peruvian individuals in our study.

**S5 Fig. a)** Boxplots showing the relationship of NHW GWAS-based PRS results between the case and control group. **b)** Figure showing ROC curves of logistic regression models of different models.

**S6 Fig.** Regional association plots, generated using LocusZoom.js[1], for the two genome-wide significant and two suggestive loci based on Model 1: **a)** *NFASC* on chromosome 1, **b)** *STK32A* on chromosome 5, **c)** *RP11-663N22.1* on chromosome 17, and **d)** *APOE* on chromosome 19. Each regional plot labels the SNPs with the lowest P value at each locus, depicting them as purple diamonds. The triangles represent individual SNPs, and the colors of the triangles indicate LD with the SNP with the lowest P value, based on the AMR reference population. The right-hand y-axis of each regional plot displays the recombination rate, marked by blue vertical lines.

**S1 Table.** The credible sets generated by CARMA.

**S2 Table.** Single-variant testing results for Known AD variants in Model 1.

**S3 Table.** Single-variant testing results for Known AD variants in Model 2.

**S4 Table.** Nominal significant genes as a result of rare variant gene-based testing for Model 1 (adjusted for sex, age, and first 4 PCs as fixed effects and GRM as a random effect), and Model 2 (also adjusted for APOE-ε4 allele dosage).

**S5 Table.** Local ancestry analysis results of the *NFASC*, *APOE*, two novel suggestive, and replicated Known-AD loci. According to the results of the ancestral dosage extraction of the lead marker in each genome-wide significant region based on 4 populations, the allele frequency is given in percent, and the allele count is given in parentheses.

### Supplementary Methods

**S1 Method.** Functional analysis

These data were obtained from three ongoing studies in Alzheimer disease. (RF1AG059018 (J. Vance PI) and U01AG072579 (J. Vance, D. Dykxhoorn, A. Griswold and J. Young MPIs) from the National Institutes on Aging of NIH, and 21A18 (K. Celis PI) grant from the Florida Department of Health Ed and Ethel Moore Alzheimer's Disease Research Program

*iPSC culture:* The iPSC lines used in this study were cultured in StemFlex™ medium (ThermoFisher Scientific) on recombinant human vitronectin (ThermoFisher Scientific) coated tissue culture plates. The cells were passaged when they achieved ~80% confluence by treatment with Accutase™ cell detachment solution (StemCell Technologies).

*Neuronal and Astrocyte differentiation:* Neurons and Astrocytes were differentiated from neuronal progenitor cells derived using the STEMdiff™ SMADi Neural Induction Kit (StemCell Technologies) using the Embryoid body (EB) approach according to a recent publication[2]. On day 12 following EB formation, STEMdiff™ Neural Rosette Selection Reagent (StemCell Technologies) was used to select the neural rosette and Neural Progenitor Cells (NPCs) expanded on Matrigel® Basement Membrane Matrix (Corning) coated plates. The NPCs were expanded before being transferred to STEMdiff™ Forebrain Neuron Differentiation Medium (StemCell Technologies) for 6 days to initiate cortical neuron differentiation.

*Forebrain neuron differentiation:* On day 25, the young neurons were passaged onto Matrigel® coated plates in STEMdiff™ Forebrain Neuron Maturation Medium (StemCell Technologies) and grown at 37°C with 5% $CO_2$. The cortical neurons were grown until day X when they were harvested for RNAseq and Hi-C analysis.

Astrocyte differentiation: For astrocyte differentiation, the media on day 13 NPCs was replaced with

STEMdiff™ Astrocyte Differentiation Medium (StemCell Technologies) for 21 days with full media changes for the first7 days followed by media changes every three days for the remaining time. The cells were grown at 37$^{o}$C with 5% $CO_2$ and the cells were passaged using Accutase™ when they reached confluence. On day 33, the media was replaced with STEMdiff™ Astrocyte Maturation Medium (StemCell Technologies) in which the cells were cultured until they were harvested on day 60 for Hi-C analysis.

*Oligodendrocyte-enriched spheroid differentiation:* Oligodendrocyte-enriched spheroids were differentiated according to the protocol of Douvaras and Fossati[3]. This approach gives a complex culture of spheroids that contain oligodendrocyte precursor cells and mature (MBP+ cells) as well as, different astrocyte and neuronal populations. The cells were harvested on day 75 for bulk Hi-C analysis.

*Microglia differentiation:* The microglia were differentiated from iPSC-derived hematopoietic progenitor cells using the STEMdiff™ Hematopoietic Kit according to the manufacturer's protocol (StemCell Technologies). On day 12, the hematopoietic progenitor cells were harvested and microglial development was induced by culturing in STEMdiff™ Microglia Differentiation medium (StemCell Technologies) for 12 days on Matrigel® (Corning) coated plates. On day 24 from the start of microglia differentiation, the cells were collected, centrifuged at 300 x g for 5 min, and replated on fresh Matrigel®-coated tissue culture plates in STEMdiff™ Microglia maturation medium. The cells were harvested on day 26 from the start of microglial differentiation for Hi-C analysis.

*Hi-C Experiment:* Hi-C analysis was performed on iPSC-derived astrocytes, microglial, oligodendrocyte-enriched spheroids, and forebrain neurons. In situ Hi-C libraries based on the protocol from Rao et al[4]. In brief, 2~5 million cells were crosslinked with 1% formaldehyde and stored at -80o until needed. Nuclei were permeabilized and DNA digested with Mbo I and the DNA ends were filled in and labelled with biotin-14-dATP

(Active Motif) before proximity ligation. Crosslinks were reversed and ligated DNA was purified, and sheared (Covaris LE220 (Covaris, Woburn, MA)) and fragments in the range of 300-500 bp were enriched using the AmPure XP beads (Beckman Coulter). Ligation junctions were pulled down with streptavidin beads and libraries prepared for sequencing on the Illumina Novaseq 6000. For each library, 450~550 million paired-end reads of 150 bp length were obtained. Sequencing data were processed using BWA [5] to map each read end separately to hg19 reference genomes. Duplicate and non-uniquely mapped reads were removed. For each library, over 270 million non-redundant, uniquely mapped, paired reads were used for further analysis. Contact matrices were generated at base pair delimited resolutions of 500 kb, 50 kb, and 5 kb. The Directionality Index (DI) value of 40kb bins was used to call the topological associated domain (TAD). For robust enhancer-promoter interaction mapping, the HiCorr pipeline was used to correct Hi-C bias at sub-TAD level to identify Hi-C loops[6].

Hi-C analysis was performed on iPSC-derived astrocytes, microglial, oligodendrocyte-enriched spheroids, and forebrain neurons. In situ Hi-C libraries based on the protocol from Rao et al. [4] For each library, 450~550 million paired-end reads of 150 bp length were obtained. For each library, over 270 million non-redundant, uniquely mapped, paired reads were used for further analysis. Contact matrices were generated at base pair delimited resolutions of 500 kb, 50 kb, and 5 kb. The Directionality Index (DI) value of 40kb bins was used to call the topological associated domain (TAD). For robust enhancer-promoter interaction mapping, the HiCorr pipeline was used to correct Hi-C bias at sub-TAD level to identify Hi-C loops[6].

### Supplementary Figures

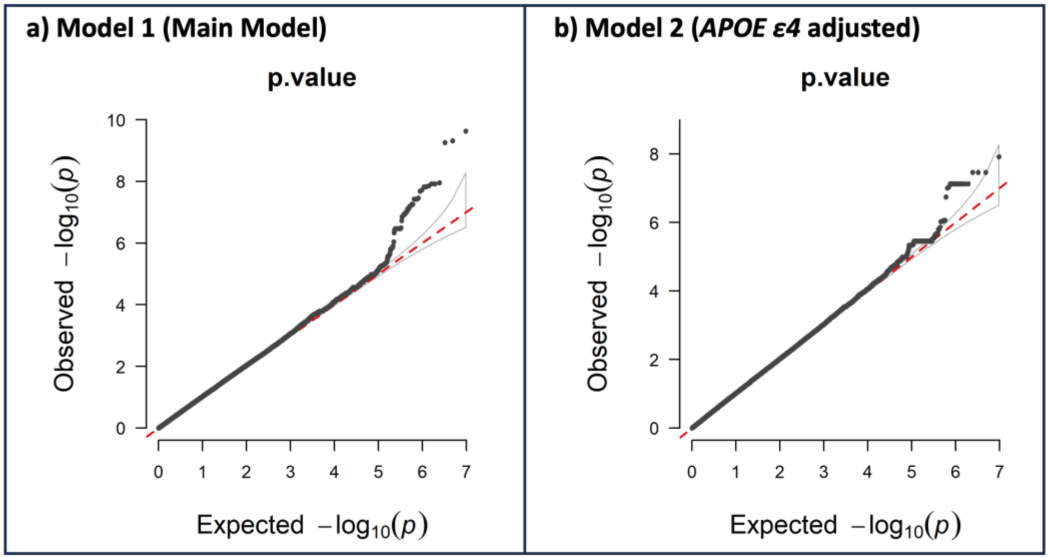


**S1 Fig.** Quantile-quantile plots show the deviation observed from expected p-values of single variant association analysis.

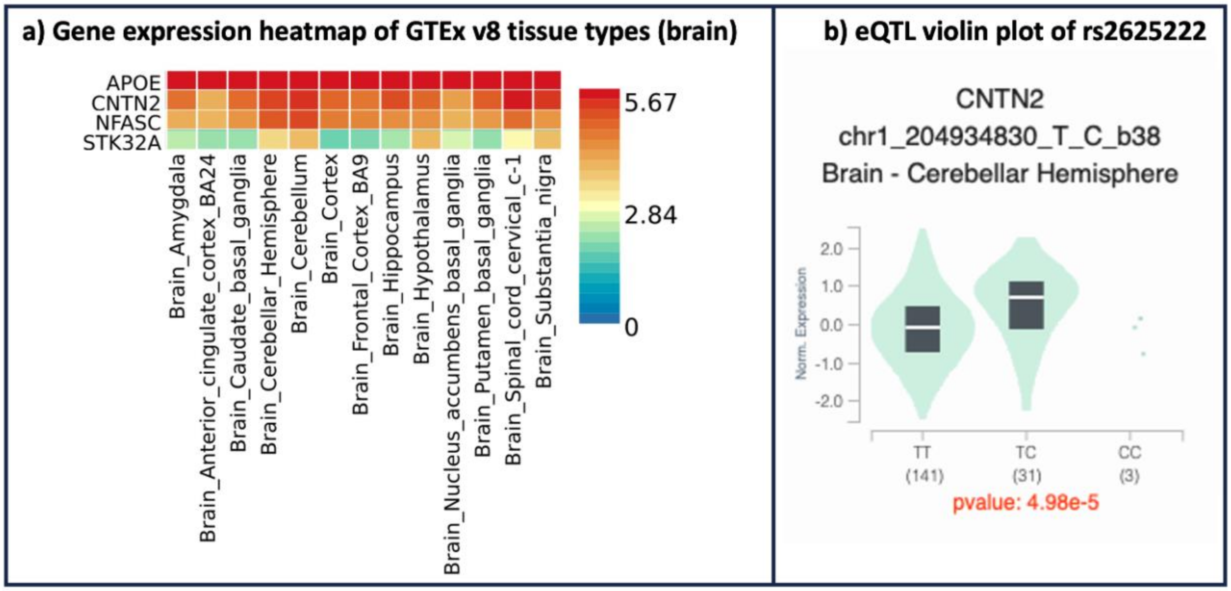


**S2 Fig. a)** Gene expression heatmap of eQTL mapped genes at $P \leq 1 \times 10^{-6}$. **b)** Single tissue (Brain - Cerebellar Hemisphere) eQTL violin plot of rs2625222 for *CNTN2* expression.

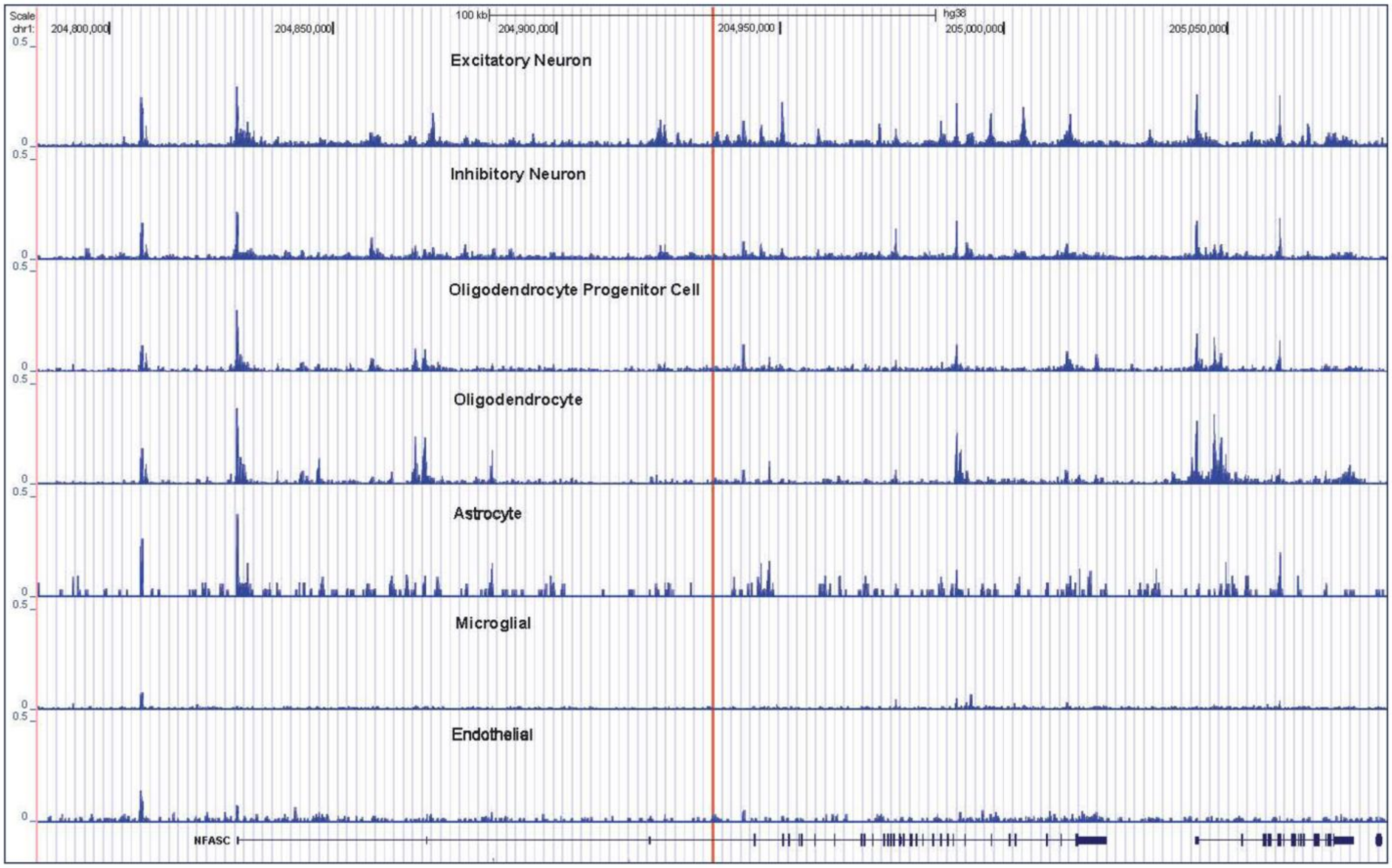


**S3 Fig.** Chromatin accessibility analysis of the region surrounding rs2625222 in frontal cortex. Visualization of chromatin accessibility by ATAC-seq peaks in the frontal cortex. rs2625222 is indicated as the red vertical line.

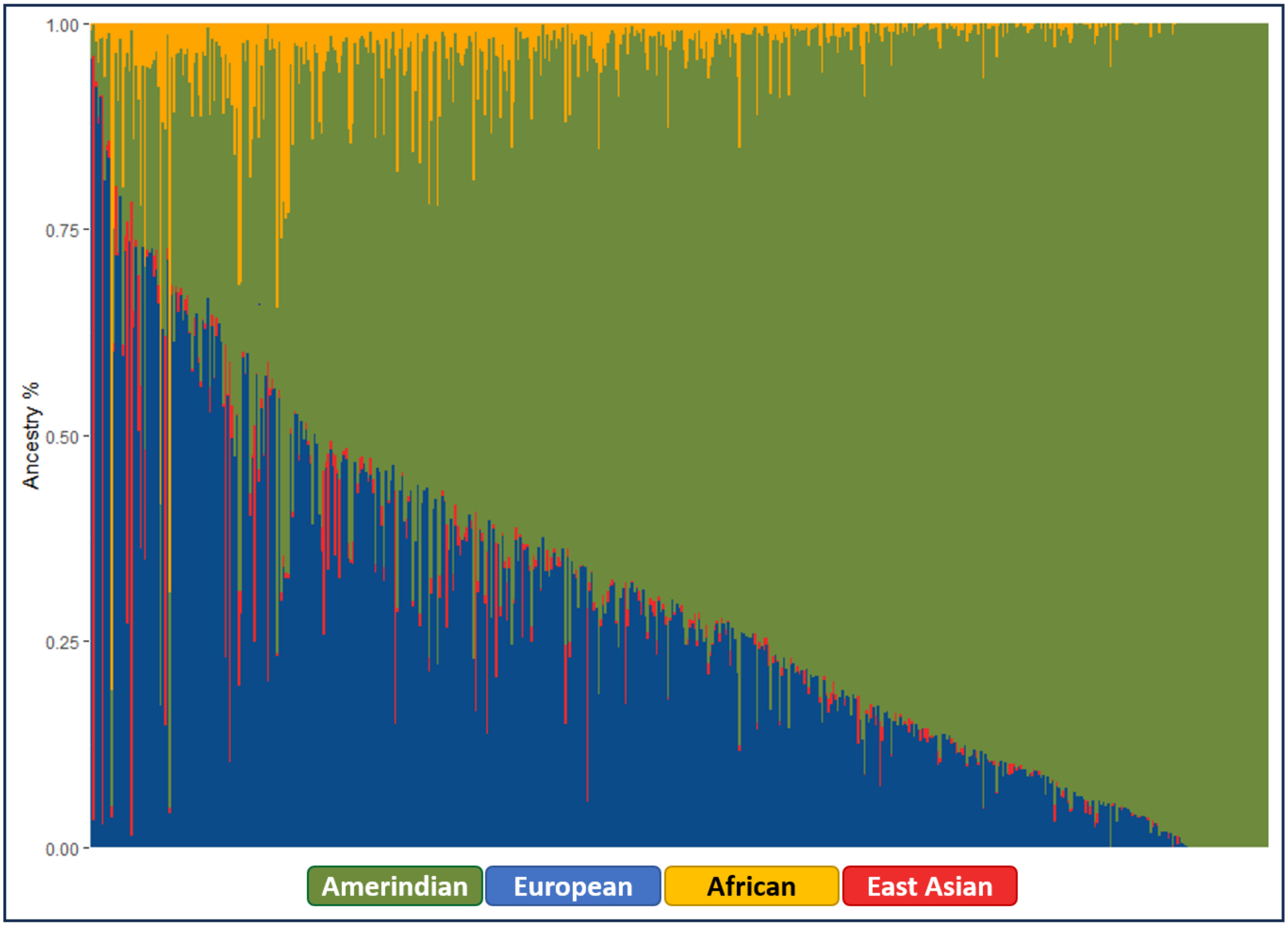


**S4 Fig.** The global ancestry bar plot of the 4-way admixed Peruvian individuals in our study.

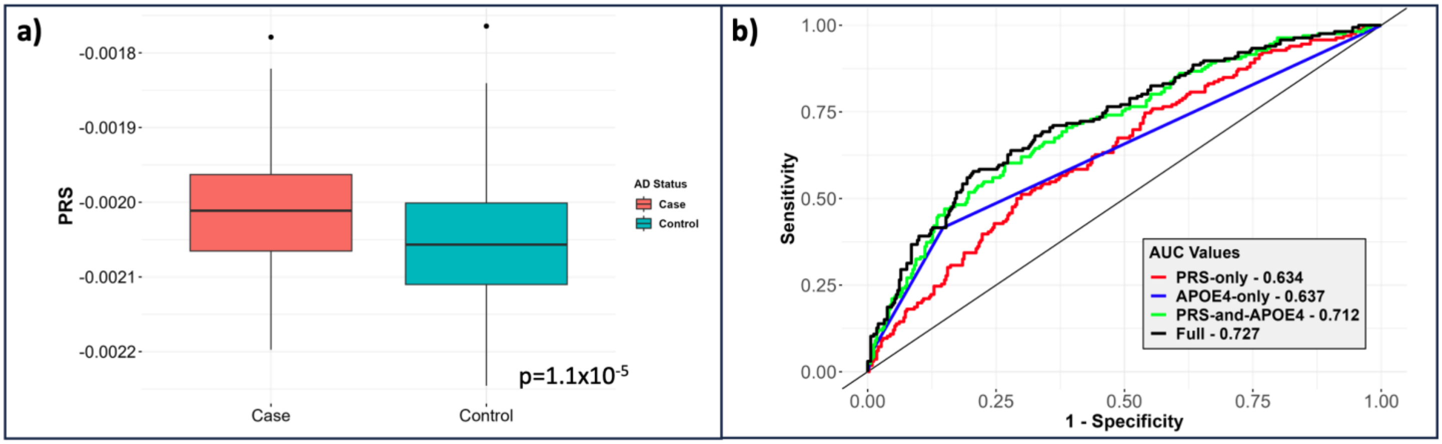


**S5 Fig. a)** Boxplots showing the relationship of NHW GWAS-based PRS results between the case and control group. **b)** Figure showing ROC curves of logistic regression models of different models.

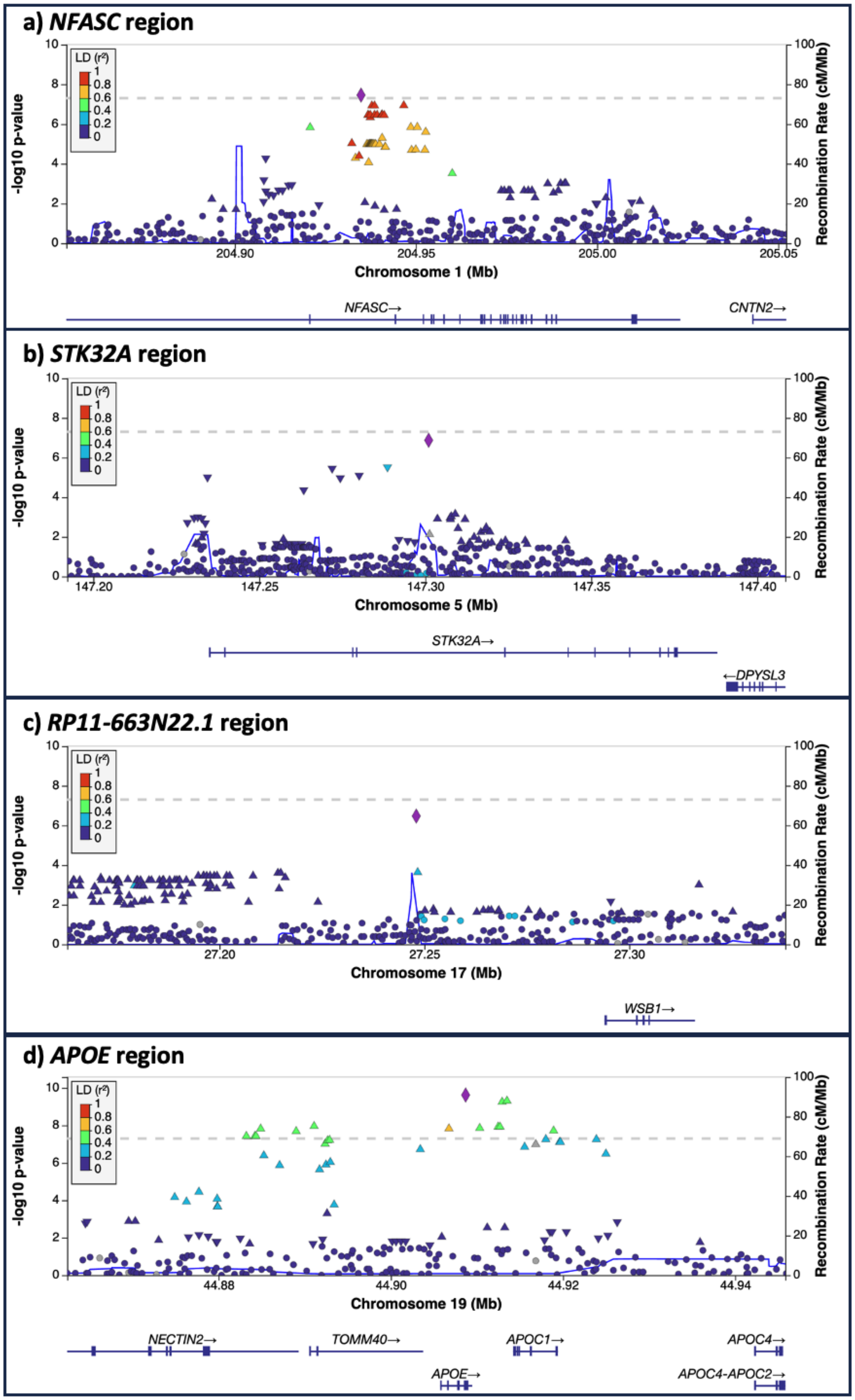


**S6 Fig.** Regional association plots, generated using LocusZoom.js[1], for the two genome-wide significant and two suggestive loci based on Model 1: **a)** *NFASC* on chromosome 1, **b)** *STK32A* on chromosome 5, **c)** *RP11-663N22.1* on chromosome 17, and **d)** *APOE* on chromosome 19. Each regional plot labels the SNPs with the lowest P value at each locus, depicting them as purple diamonds. The triangles represent individual SNPs, and the colors of the triangles indicate LD with the SNP with the lowest P value, based on the AMR reference population. The right-hand y-axis of each regional plot displays the recombination rate, marked by blue vertical lines.

### Supplementary Tables

**S1 Table.** The credible sets generated by CARMA.

| Marker | Allele1 | Allele2 | BETA | SE | p.value | SEI | CADD | PIP |
|---|---|---|---|---|---|---|---|---|
| ***NFASC* region (Model 1)** | | | | | | | | |
| chr1:204934830 | T | C | 1.97 | 0.357 | 1.58e-08 | 0.0299 | 10.9 | 0.39 |
| chr1:204937811 | G | T | 1.86 | 0.35 | 5.43e-08 | 0.0463 | 3.48 | 0.0969 |
| chr1:204946629 | G | A | 1.86 | 0.349 | 5.25e-08 | 0.167 | 2.63 | 0.0935 |
| chr1:204938561 | A | T | 1.86 | 0.35 | 5.43e-08 | 0.0393 | 1.4 | 0.093 |
| chr1:204940601 | C | T | 1.69 | 0.332 | 1.76e-07 | 0.0624 | 9.59 | 0.0349 |
| chr1:204941233 | T | A | 1.75 | 0.343 | 1.65e-07 | 0.175 | 6.1 | 0.0339 |
| chr1:204936780 | A | G | 1.69 | 0.332 | 1.76e-07 | 0.201 | 6.7 | 0.0328 |
| chr1:204938554 | A | G | 1.69 | 0.332 | 1.76e-07 | 0.105 | 5.31 | 0.0321 |
| chr1:204940644 | T | C | 1.69 | 0.332 | 1.76e-07 | 0.255 | 4.01 | 0.031 |
| chr1:204937385 | A | G | 1.69 | 0.332 | 1.76e-07 | 0.0259 | 2.71 | 0.0306 |
| chr1:204939288 | A | G | 1.69 | 0.332 | 1.76e-07 | 0.0658 | 4.4 | 0.0201 |
| chr1:204940588 | CCTT | C | 1.69 | 0.332 | 1.76e-07 | 0.573 | 3.44 | 0.0192 |
| chr1:204936792 | G | A | 1.69 | 0.332 | 1.76e-07 | 0.186 | 1.82 | 0.019 |
| chr1:204940580 | G | A | 1.69 | 0.332 | 1.76e-07 | 0.226 | 0.563 | 0.0185 |
| chr1:204937371 | T | C | 1.67 | 0.33 | 2.17e-07 | 0.0187 | 5.17 | 0.0168 |
| ***APOE* region (Model 1)** | | | | | | | | |
| chr19:44908684 | T | C | 1.13 | 0.192 | 1.96e-09 | 0.397 | 16.6 | 0.821 |
| chr19:44912921 | G | T | 1.06 | 0.182 | 2.95e-09 | 0.101 | 11.4 | 0.155 |
| ***NFASC* region (Model 2)** | | | | | | | | |
| chr1:204934830 | T | C | 2.06 | 0.366 | 9.04e-09 | 0.0299 | 10.9 | 0.281 |
| chr1:204937811 | G | T | 1.96 | 0.359 | 2.58e-08 | 0.0463 | 3.48 | 0.094 |
| chr1:204938561 | A | T | 1.96 | 0.359 | 2.58e-08 | 0.0393 | 1.4 | 0.0917 |
| chr1:204946629 | G | A | 1.95 | 0.359 | 2.65e-08 | 0.167 | 2.63 | 0.0878 |
| chr1:204920781 | G | A | 2.55 | 0.489 | 9.05e-08 | 0.161 | 0.975 | 0.0744 |
| chr1:204940601 | C | T | 1.81 | 0.342 | 6.16e-08 | 0.0624 | 9.59 | 0.0435 |
| chr1:204938554 | A | G | 1.81 | 0.342 | 6.16e-08 | 0.105 | 5.31 | 0.0413 |
| chr1:204939288 | A | G | 1.81 | 0.342 | 6.16e-08 | 0.0658 | 4.4 | 0.0409 |
| chr1:204940644 | T | C | 1.81 | 0.342 | 6.16e-08 | 0.255 | 4.01 | 0.0404 |
| chr1:204940588 | CCTT | C | 1.81 | 0.342 | 6.16e-08 | 0.573 | 3.44 | 0.0397 |
| chr1:204941233 | T | A | 1.86 | 0.352 | 6.95e-08 | 0.175 | 6.1 | 0.0369 |
| chr1:204937371 | T | C | 1.78 | 0.34 | 7.53e-08 | 0.0187 | 5.17 | 0.0343 |
| chr1:204936780 | A | G | 1.81 | 0.342 | 6.16e-08 | 0.201 | 6.7 | 0.0277 |
| chr1:204937385 | A | G | 1.81 | 0.342 | 6.16e-08 | 0.0259 | 2.71 | 0.0266 |
| ***RP5-1121H13.4* region (Model 2)** | | | | | | | | |
| chr20:41869905 | A | C | 1.29 | 0.262 | 4.35e-07 | 0.0213 | 1.2 | 0.912 |
| chr20:41870060 | C | T | 0.929 | 0.228 | 2.26e-05 | 0.019 | 1.22 | 0.0218 |
| chr20:41859849 | G | A | 1.03 | 0.265 | 5.33e-05 | 0.0344 | 0.342 | 0.0135 |
| chr20:41860676 | A | T | 1.03 | 0.265 | 5.33e-05 | 0.0114 | 0.727 | 0.0127 |

**S2 Table.** Single-variant testing results for Known AD variants[7] in Model 1.

| Marker | dbSNP | Closest gene | Reference Allele | Effect Allele | BETA | SE | p.value |
|---|---|---|---|---|---|---|---|
| chr1:207577223 | rs679515 | *CR1* | T | C | 0.090186 | 0.312689 | 0.773024 |
| chr2:9558882 | rs72777026 | *ADAM17* | A | G | -0.07716 | 0.216583 | 0.721659 |
| chr2:37304796 | rs17020490 | *PRKD3* | T | C | 0.13947 | 0.14289 | 0.329034 |
| chr2:127135234 | rs6733839 | *BIN1* | C | T | 0.147699 | 0.138726 | 0.28702 |
| chr2:202878716 | rs139643391 | *WDR12* | TC | T | 0.027675 | 0.32489 | 0.932117 |
| chr2:233117202 | rs10933431 | *INPP5D* | G | C | 0.151004 | 0.12944 | 0.243374 |
| chr3:155069722 | rs16824536 | *MME* | G | A | -0.29517 | 0.397904 | 0.458198 |
| chr4:993555 | rs3822030 | *IDUA* | GAGTT | G | -0.15007 | 0.454969 | 0.741516 |
| chr4:993555 | rs3822030 | *IDUA* | G | T | -0.0137 | 0.140222 | 0.922188 |
| chr4:11023507 | rs6846529 | *CLNK* | C | T | -0.14135 | 0.142332 | 0.320672 |
| chr4:40197226 | rs2245466 | *RHOH* | G | C | -0.17441 | 0.137912 | 0.206001 |
| chr5:14724304 | rs112403360 | *ANKH* | T | A | -0.05634 | 0.504767 | 0.911134 |
| chr5:86927378 | rs62374257 | *COX7C* | T | C | 0.017615 | 0.156185 | 0.910201 |
| chr5:151052827 | rs871269 | *TNIP1* | C | T | -0.06563 | 0.143766 | 0.648044 |
| chr5:180201150 | rs113706587 | *RASGEF1C* | G | A | 0.207431 | 0.354963 | 0.558968 |
| chr6:32615322 | rs6605556 | *HLA-DQA1* | A | G | 0.020947 | 0.155176 | 0.89262 |
| chr6:41036354 | rs10947943 | *UNC5CL* | G | A | -0.16012 | 0.159052 | 0.314071 |
| chr6:41181270 | rs60755019 | *TREML2* | A | G | 0.751677 | 0.291447 | 0.004953 |
| chr6:47517390 | rs7767350 | *CD2AP* | C | T | 0.250131 | 0.170242 | 0.141759 |
| chr6:114291731 | rs785129 | *HS3ST5* | T | C | -0.00884 | 0.143694 | 0.950937 |
| chr7:7817263 | rs6943429 | *UMAD1* | T | C | -0.11629 | 0.134666 | 0.387838 |
| chr7:8204382 | rs10952097 | *ICA1* | T | C | 0.051712 | 0.285792 | 0.856413 |
| chr7:12229967 | rs13237518 | *TMEM106B* | C | A | 0.068153 | 0.13644 | 0.61742 |
| chr7:28129126 | rs1160871 | *JAZF1* | GTCTT | G | -0.11352 | 0.152474 | 0.456582 |
| chr7:37844191 | rs6966331 | *EPDR1* | T | C | 0.120711 | 0.144991 | 0.405106 |
| chr7:54873635 | rs76928645 | *SEC61G* | C | T | -0.71432 | 0.388414 | 0.065905 |
| chr7:100334426 | rs7384878 | *SPDYE3* | C | T | -0.09714 | 0.156311 | 0.5343 |
| chr7:143413669 | rs11771145 | *EPHA1* | G | A | 0.138904 | 0.150518 | 0.356089 |
| chr8:11844613 | rs1065712 | *CTSB* | G | C | -0.39789 | 0.69533 | 0.567164 |
| chr8:27362470 | rs73223431 | *PTK2B* | C | T | -0.1099 | 0.180447 | 0.542494 |
| chr8:27607795 | rs11787077 | *CLU* | T | C | 0.315615 | 0.158875 | 0.023486 |
| chr9:104903697 | rs1800978 | *ABCA1* | C | G | 0.209195 | 0.164655 | 0.203904 |
| chr10:11676714 | rs7912495 | *USP6NL* | A | G | 0.244421 | 0.141768 | 0.084691 |
| chr10:60025170 | rs7068231 | *ANK3* | T | G | -0.0032 | 0.148003 | 0.982734 |
| chr10:80494228 | rs6586028 | *TSPAN14* | C | T | -0.21756 | 0.231551 | 0.347427 |
| chr10:122413396 | rs7908662 | *PLEKHA1* | A | G | -0.26485 | 0.155857 | 0.089263 |
| chr11:47370397 | rs10437655 | *SPI1* | G | A | 0.235667 | 0.18197 | 0.19529 |
| chr11:60254475 | rs1582763 | *MS4A4A* | G | A | 0.051788 | 0.140046 | 0.711537 |
| chr11:86157598 | rs3851179 | *EED* | T | C | 0.185511 | 0.138503 | 0.180439 |
| chr11:121564878 | rs11218343 | *SORL1* | T | C | 0.213409 | 0.295344 | 0.469939 |
| chr12:113281983 | rs6489896 | *TPCN1* | T | C | -0.26986 | 0.279957 | 0.335084 |
| chr14:52924962 | rs17125924 | *FERMT2* | A | G | -0.13793 | 0.168276 | 0.412393 |

| | | | | | | | |
|---|---|---|---|---|---|---|---|
| chr14:92464917 | rs7401792 | *SLC24A4* | G | A | -0.07211 | 0.14226 | 0.612251 |
| chr15:50701814 | rs8025980 | *SPPL2A* | A | G | -0.17831 | 0.144804 | 0.218187 |
| chr15:58764824 | rs602602 | *MINDY2* | T | A | 0.025656 | 0.141257 | 0.855878 |
| chr15:63277703 | rs117618017 | *APH1B* | C | T | -0.36769 | 0.378348 | 0.331134 |
| chr15:64131307 | rs3848143 | *SNX1* | G | A | 0.283917 | 0.164244 | 0.041938 |
| chr15:78936857 | rs12592898 | *CTSH* | A | G | -0.21612 | 0.254799 | 0.396324 |
| chr16:30010081 | rs1140239 | *DOC2A* | C | T | 0.027287 | 0.155168 | 0.860409 |
| chr16:31111250 | rs889555 | *BCKDK* | C | T | 0.009169 | 0.13155 | 0.944434 |
| chr16:70660097 | rs4985556 | *IL34* | C | A | 0.199543 | 0.319828 | 0.53269 |
| chr16:79574511 | rs450674 | *MAF* | T | C | 0.03964 | 0.142274 | 0.780537 |
| chr16:81739398 | rs12446759 | *PLCG2* | G | A | 0.18422 | 0.132621 | 0.164811 |
| chr16:86420604 | rs16941239 | *FOXF1* | T | A | 0.006816 | 0.332584 | 0.983649 |
| chr16:90103687 | rs56407236 | *PRDM7* | G | A | 0.387412 | 0.240557 | 0.107294 |
| chr17:1728046 | rs35048651 | *WDR81* | TGAG | T | -0.0085 | 0.153756 | 0.955897 |
| chr17:5233752 | rs7225151 | *SCIMP* | G | A | 0.237893 | 0.36696 | 0.516804 |
| chr17:18156140 | rs2242595 | *MYO15A* | G | A | -0.0932 | 0.140499 | 0.507103 |
| chr17:44352876 | rs5848 | *GRN* | C | T | -0.2026 | 0.179441 | 0.258871 |
| chr17:46779275 | rs199515 | *WNT3* | G | C | -0.00483 | 0.243448 | 0.984158 |
| chr17:58332680 | rs2526377 | *TSPOAP1* | A | G | -0.0122 | 0.136806 | 0.928932 |
| chr17:63471557 | rs4277405 | *ACE* | C | T | 0.089163 | 0.156886 | 0.569813 |
| chr19:1050875 | rs12151021 | *ABCA7* | A | G | -0.14666 | 0.205262 | 0.474911 |
| chr19:1854254 | rs149080927 | *KLF16* | G | GC | 0.002179 | 0.148954 | 0.988328 |
| chr19:49950060 | rs9304690 | *SIGLEC11* | C | T | -0.01715 | 0.229308 | 0.940391 |
| chr19:54267597 | rs587709 | *LILRB2* | C | T | -0.03662 | 0.144975 | 0.800595 |
| chr20:413334 | rs1358782 | *RBCK1* | A | G | -0.04488 | 0.218711 | 0.837398 |
| chr20:56423488 | rs6014724 | *CASS4* | A | G | 0.13687 | 0.162643 | 0.400047 |
| chr20:63743088 | rs6742 | *SLC2A4RG* | T | C | -0.18737 | 0.232334 | 0.419976 |
| chr21:26101558 | rs2154481 | *APP* | C | T | -0.07521 | 0.158916 | 0.636001 |
| chr21:26775872 | rs2830489 | *ADAMTS1* | C | T | -0.29943 | 0.232752 | 0.198273 |

**S3 Table.** Single-variant testing results for Known AD variants[7] in Model 2.

| Marker | dbSNP | Closest gene | Reference Allele | Effect Allele | BETA | SE | p.value |
|---|---|---|---|---|---|---|---|
| chr1:207577223 | rs679515 | *CR1* | T | C | 0.121636 | 0.326167 | 0.709204 |
| chr2:9558882 | rs72777026 | *ADAM17* | A | G | -0.10337 | 0.224789 | 0.645627 |
| chr2:37304796 | rs17020490 | *PRKD3* | T | C | 0.09812 | 0.149653 | 0.51205 |
| chr2:127135234 | rs6733839 | *BIN1* | C | T | 0.052117 | 0.145041 | 0.719355 |
| chr2:202878716 | rs139643391 | *WDR12* | TC | T | 0.000825 | 0.337738 | 0.99805 |
| chr2:233117202 | rs10933431 | *INPP5D* | G | C | 0.079768 | 0.136395 | 0.558662 |
| chr3:155069722 | rs16824536 | *MME* | G | A | -0.17995 | 0.421057 | 0.669107 |
| chr4:993555 | rs3822030 | *IDUA* | GAGTT | G | 0.071477 | 0.482907 | 0.882332 |
| chr4:993555 | rs3822030 | *IDUA* | G | T | -0.08028 | 0.147584 | 0.586453 |
| chr4:11023507 | rs6846529 | *CLNK* | C | T | -0.15773 | 0.148591 | 0.288464 |
| chr4:40197226 | rs2245466 | *RHOH* | G | C | -0.22093 | 0.143284 | 0.12309 |
| chr5:14724304 | rs112403360 | *ANKH* | T | A | 0.068439 | 0.53262 | 0.897758 |
| chr5:86927378 | rs62374257 | *COX7C* | T | C | 0.01693 | 0.163815 | 0.917686 |
| chr5:151052827 | rs871269 | *TNIP1* | C | T | -0.11622 | 0.150004 | 0.438467 |
| chr5:180201150 | rs113706587 | *RASGEF1C* | G | A | 0.220363 | 0.372346 | 0.553969 |
| chr6:32615322 | rs6605556 | *HLA-DQA1* | A | G | 0.006044 | 0.161325 | 0.970113 |
| chr6:41036354 | rs10947943 | *UNC5CL* | G | A | -0.20723 | 0.166061 | 0.212056 |
| chr6:41181270 | rs60755019 | *TREML2* | A | G | 0.790892 | 0.300138 | 0.004206 |
| chr6:47517390 | rs7767350 | *CD2AP* | C | T | 0.124321 | 0.179364 | 0.488232 |
| chr6:114291731 | rs785129 | *HS3ST5* | T | C | 0.006203 | 0.150926 | 0.967218 |
| chr7:7817263 | rs6943429 | *UMAD1* | T | C | -0.06374 | 0.140832 | 0.650851 |
| chr7:8204382 | rs10952097 | *ICA1* | T | C | 0.072793 | 0.294616 | 0.804848 |
| chr7:12229967 | rs13237518 | *TMEM106B* | C | A | 0.060751 | 0.143752 | 0.672584 |
| chr7:28129126 | rs1160871 | *JAZF1* | GTCTT | G | -0.18157 | 0.159922 | 0.256225 |
| chr7:37844191 | rs6966331 | *EPDR1* | T | C | 0.059074 | 0.152116 | 0.697758 |
| chr7:54873635 | rs76928645 | *SEC61G* | C | T | -0.63668 | 0.40413 | 0.115154 |
| chr7:100334426 | rs7384878 | *SPDYE3* | C | T | -0.17534 | 0.164271 | 0.285808 |
| chr7:143413669 | rs11771145 | *EPHA1* | G | A | 0.041041 | 0.157867 | 0.794884 |
| chr8:11844613 | rs1065712 | *CTSB* | G | C | -0.49034 | 0.716557 | 0.493789 |
| chr8:27362470 | rs73223431 | *PTK2B* | C | T | -0.10363 | 0.188449 | 0.582381 |
| chr8:27607795 | rs11787077 | *CLU* | T | C | 0.299654 | 0.170311 | 0.03925 |
| chr9:104903697 | rs1800978 | *ABCA1* | C | G | 0.139409 | 0.172865 | 0.419975 |
| chr10:11676714 | rs7912495 | *USP6NL* | A | G | 0.278828 | 0.148954 | 0.061219 |
| chr10:60025170 | rs7068231 | *ANK3* | T | G | -0.03312 | 0.156338 | 0.832214 |
| chr10:80494228 | rs6586028 | *TSPAN14* | C | T | -0.11439 | 0.242233 | 0.636774 |
| chr10:122413396 | rs7908662 | *PLEKHA1* | A | G | -0.3161 | 0.163179 | 0.05273 |
| chr11:47370397 | rs10437655 | *SPI1* | G | A | 0.162436 | 0.192124 | 0.397846 |
| chr11:60254475 | rs1582763 | *MS4A4A* | G | A | -0.04105 | 0.146122 | 0.778752 |
| chr11:86157598 | rs3851179 | *EED* | T | C | 0.183128 | 0.145901 | 0.209423 |
| chr11:121564878 | rs11218343 | *SORL1* | T | C | 0.125631 | 0.308432 | 0.683772 |
| chr12:113281983 | rs6489896 | *TPCN1* | T | C | -0.35422 | 0.288865 | 0.22011 |

| | | | | | | | |
|---|---|---|---|---|---|---|---|
| chr14:52924962 | rs17125924 | *FERMT2* | A | G | -0.11821 | 0.17501 | 0.499409 |
| chr14:92464917 | rs7401792 | *SLC24A4* | G | A | -0.0402 | 0.148995 | 0.787299 |
| chr15:50701814 | rs8025980 | *SPPL2A* | A | G | -0.1794 | 0.152556 | 0.239609 |
| chr15:58764824 | rs602602 | *MINDY2* | T | A | 0.109399 | 0.148614 | 0.461653 |
| chr15:63277703 | rs117618017 | *APH1B* | C | T | -0.62067 | 0.403822 | 0.124297 |
| chr15:64131307 | rs3848143 | *SNX1* | G | A | 0.309829 | 0.170137 | 0.034299 |
| chr15:78936857 | rs12592898 | *CTSH* | A | G | -0.21598 | 0.26727 | 0.419038 |
| chr16:30010081 | rs1140239 | *DOC2A* | C | T | -0.02798 | 0.162408 | 0.863211 |
| chr16:31111250 | rs889555 | *BCKDK* | C | T | -0.06474 | 0.13764 | 0.638092 |
| chr16:70660097 | rs4985556 | *IL34* | C | A | 0.093596 | 0.33617 | 0.780692 |
| chr16:79574511 | rs450674 | *MAF* | T | C | 0.135758 | 0.150435 | 0.366824 |
| chr16:81739398 | rs12446759 | *PLCG2* | G | A | 0.127649 | 0.139028 | 0.358538 |
| chr16:86420604 | rs16941239 | *FOXF1* | T | A | -0.01408 | 0.346791 | 0.967615 |
| chr16:90103687 | rs56407236 | *PRDM7* | G | A | 0.381849 | 0.24946 | 0.125844 |
| chr17:1728046 | rs35048651 | *WDR81* | TGAG | T | -0.06116 | 0.160698 | 0.703488 |
| chr17:5233752 | rs7225151 | *SCIMP* | G | A | 0.461065 | 0.391043 | 0.238372 |
| chr17:18156140 | rs2242595 | *MYO15A* | G | A | -0.0638 | 0.146054 | 0.662218 |
| chr17:44352876 | rs5848 | *GRN* | C | T | -0.20479 | 0.188801 | 0.278062 |
| chr17:46779275 | rs199515 | *WNT3* | G | C | -0.02159 | 0.25615 | 0.932823 |
| chr17:58332680 | rs2526377 | *TSPOAP1* | A | G | 0.05613 | 0.143896 | 0.696483 |
| chr17:63471557 | rs4277405 | *ACE* | C | T | 0.1301 | 0.16459 | 0.429267 |
| chr19:1050875 | rs12151021 | *ABCA7* | A | G | -0.1593 | 0.215137 | 0.45901 |
| chr19:1854254 | rs149080927 | *KLF16* | G | GC | 0.035443 | 0.156106 | 0.82039 |
| chr19:49950060 | rs9304690 | *SIGLEC11* | C | T | -0.08222 | 0.239335 | 0.731196 |
| chr19:54267597 | rs587709 | *LILRB2* | C | T | -0.04853 | 0.15208 | 0.749632 |
| chr20:413334 | rs1358782 | *RBCK1* | A | G | -0.19371 | 0.23138 | 0.402494 |
| chr20:56423488 | rs6014724 | *CASS4* | A | G | 0.152507 | 0.169085 | 0.367082 |
| chr20:63743088 | rs6742 | *SLC2A4RG* | T | C | -0.14795 | 0.241213 | 0.539633 |
| chr21:26101558 | rs2154481 | *APP* | C | T | -0.00585 | 0.167226 | 0.972085 |
| chr21:26775872 | rs2830489 | *ADAMTS1* | C | T | -0.263 | 0.243432 | 0.27998 |

**S4 Table.** Nominal significant genes as a result of rare variant gene-based testing for Model 1 (adjusted for sex, age, and first 4 PCs as fixed effects and GRM as a random effect), and Model 2 (also adjusted for *APOE-ε4* allele dosage).

| | | group | n.variants | freq.mean | B.score | B.var | B.pval | S.pval | O.pval | O.minp | O.minp.rho | O.pval. Bonferroni |
|---|---|---|---|---|---|---|---|---|---|---|---|---|
| **Model 1** | **CADD0** | *ACE* | 57 | 0.0030 | 776.23 | 106005.89 | 0.017 | 0.070 | 0.023 | 0.017 | 1 | 1 |
| | | *CTSH* | 66 | 0.0084 | -850.74 | 144257.18 | 0.025 | 0.090 | 0.032 | 0.025 | 1 | 1 |
| | | *KLF16* | 20 | 0.0121 | -236.21 | 17066.19 | 0.071 | 0.027 | 0.034 | 0.027 | 0 | 1 |
| | | *PICALM* | 540 | 0.0034 | 1179.99 | 2878262.07 | 0.487 | 0.020 | 0.037 | 0.020 | 0 | 1 |
| | | *MME* | 225 | 0.0115 | -2556.76 | 1370660.23 | 0.029 | 0.256 | 0.038 | 0.029 | 1 | 1 |
| | | *WDR81* | 42 | 0.0025 | 166.18 | 32088.82 | 0.354 | 0.029 | 0.048 | 0.029 | 0 | 1 |
| | **CADD10** | *TREM2* | 5 | 0.0119 | 70.77 | 2042.68 | 0.117 | 0.013 | 0.024 | 0.013 | 0 | 1 |
| | | *PICALM* | 50 | 0.0035 | 156.85 | 36033.08 | 0.409 | 0.023 | 0.043 | 0.023 | 0 | 1 |
| | | *CASS4* | 6 | 0.0043 | 89.84 | 1718.47 | 0.030 | 0.455 | 0.046 | 0.030 | 1 | 1 |
| | **CADD20** | *TREM2* | 3 | 0.0193 | 85.73 | 1815.46 | 0.044 | 0.012 | 0.021 | 0.012 | 0.09 | 1 |
| **Model 2** | **CADD0** | *MME* | 225 | 0.0115 | -2796.03 | 1228664.12 | 0.012 | 0.068 | 0.014 | 0.012 | 1 | 1 |
| | | *ACE* | 57 | 0.0030 | 774.74 | 95980.06 | 0.012 | 0.041 | 0.016 | 0.012 | 1 | 1 |
| | | *SNX1* | 161 | 0.0060 | 1127.44 | 1165414.66 | 0.296 | 0.022 | 0.034 | 0.022 | 0 | 1 |
| | | *KLF16* | 20 | 0.0121 | -220.83 | 15730.76 | 0.078 | 0.035 | 0.044 | 0.035 | 0 | 1 |
| | **CADD10** | *CASS4* | 6 | 0.0043 | 93.63 | 1461.20 | 0.014 | 0.301 | 0.021 | 0.014 | 1 | 1 |
| | | *TREM2* | 5 | 0.0119 | 60.26 | 1823.77 | 0.158 | 0.018 | 0.032 | 0.018 | 0 | 1 |
| | | *SLC24A4* | 28 | 0.0063 | -250.03 | 12221.24 | 0.024 | 0.150 | 0.044 | 0.024 | 1 | 1 |
| | | *PRKD3* | 18 | 0.0031 | 102.71 | 8609.26 | 0.268 | 0.027 | 0.044 | 0.027 | 0 | 1 |
| | **CADD20** | *TREM2* | 3 | 0.0193 | 72.05 | 1626.37 | 0.074 | 0.015 | 0.027 | 0.015 | 0 | 1 |

**S5 Table 5.** Local ancestry analysis results of the *NFASC*, *APOE*, two novel suggestive, and replicated Known-AD loci. According to the results of the ancestral dosage extraction of the lead marker in each genome-wide significant region based on 4 populations, the allele frequency is given in percent, and the allele count is given in parentheses.

| | | Alternative Allele Frequency (Alternative Allele Count) | | | |
|---|---|---|---|---|---|
| | | **AFR** | **AMR** | **EAS** | **EUR** |
| **rs2625222 (*NFASC*)** | **Controls** | 3.6% (1) | 0% (0) | 0% (0) | 4.2% (8) |
| | **Cases** | 29.2% (7) | 0% (0) | 0% (0) | 21.1% (23) |
| **rs4441917 (*STK32A*)** | **Controls** | 26.1% (6) | 49.5% (234) | 20% (3) | 28.2% (55) |
| | **Cases** | 21.1% (4) | 28.4% (60) | 20% (1) | 21.7% (25) |
| **rs56281491 (*RP11-663N22.1*)** | **Controls** | 36.8% (7) | 48.7% (230) | 27.8% (5) | 55.3% (109) |
| | **Cases** | 66.7% (14) | 66.4% (140) | 54.5% (6) | 64.5% (69) |
| **rs429358 (*APOE*)** | **Controls** | 19.2% (5) | 6.3% (29) | 15.4% (2) | 10.8% (22) |
| | **Cases** | 45.5% (10) | 17.5% (37) | 33.3% (1) | 34.2% (39) |
| **rs60755019 (*TREML2*)** | **Controls** | 26.9% (7) | 4.8% (22) | 0% (0) | 0% (0) |
| | **Cases** | 22.2% (4) | 11.1% (25) | 0% (0) | 1.9% (2) |
| **rs11787077 (*CLU*)** | **Controls** | 55% (11) | 39.7% (192) | 10% (1) | 38.5% (74) |
| | **Cases** | 63.6% (7) | 32.1% (75) | 50% (3) | 33.3% (33) |

#### Supplemental References


1. Boughton AP, Welch RP, Flickinger M, VandeHaar P, Taliun D, Abecasis GR, et al. LocusZoom.js: interactive and embeddable visualization of genetic association study results. Bioinformatics. 2021;37(18):3017-8. doi: 10.1093/bioinformatics/btab186. PubMed PMID: 33734315; PubMed Central PMCID: PMCPMC8479674.
2. Cukier HN, Duarte CL, Laverde-Paz MJ, Simon SA, Van Booven DJ, Miyares AT, et al. An Alzheimer's disease risk variant in TTC3 modifies the actin cytoskeleton organization and the PI3K-Akt signaling pathway in iPSC-derived forebrain neurons. bioRxiv. 2023. Epub 20230525. doi: 10.1101/2023.05.25.542316. PubMed PMID: 37292815; PubMed Central PMCID: PMCPMC10246004.
3. Douvaras P, Fossati V. Generation and isolation of oligodendrocyte progenitor cells from human pluripotent stem cells. Nat Protoc. 2015;10(8):1143-54. Epub 20150702. doi: 10.1038/nprot.2015.075. PubMed PMID: 26134954.
4. Rao SS, Huntley MH, Durand NC, Stamenova EK, Bochkov ID, Robinson JT, et al. A 3D map of the human genome at kilobase resolution reveals principles of chromatin looping. Cell. 2014;159(7):1665-80. Epub 20141211. doi: 10.1016/j.cell.2014.11.021. PubMed PMID: 25497547; PubMed Central PMCID: PMCPMC5635824.
5. Genomes Project C, Auton A, Brooks LD, Durbin RM, Garrison EP, Kang HM, et al. A global reference for human genetic variation. Nature. 2015;526(7571):68-74. doi: 10.1038/nature15393. PubMed PMID: 26432245; PubMed Central PMCID: PMCPMC4750478.
6. Lu L, Liu X, Huang WK, Giusti-Rodriguez P, Cui J, Zhang S, et al. Robust Hi-C Maps of Enhancer-Promoter Interactions Reveal the Function of Non-coding Genome in Neural Development and Diseases. Mol Cell. 2020;79(3):521-34 e15. Epub 20200626. doi: 10.1016/j.molcel.2020.06.007. PubMed PMID: 32592681; PubMed Central PMCID: PMCPMC7415676.

7. Bellenguez C, Kucukali F, Jansen IE, Kleineidam L, Moreno-Grau S, Amin N, et al. New insights into the genetic etiology of Alzheimer's disease and related dementias. Nat Genet. 2022;54(4):412-36. Epub 20220404. doi: 10.1038/s41588-022-01024-z. PubMed PMID: 35379992; PubMed Central PMCID: PMCPMC9005347.
